# Multicenter Histology Image Integration and Multiscale Deep Learning for Machine Learning-Enabled Pediatric Sarcoma Classification

**DOI:** 10.1101/2025.06.10.25328700

**Authors:** Adam Thiesen, Sergii Domanskyi, Ali Foroughi pour, Jingyan Zhang, Todd B. Sheridan, Steven B. Neuhauser, Alyssa Stetson, Katelyn Dannheim, Danielle B. Cameron, Shawn Ahn, Hao Wu, Emily R. Christison Lagay, Carol J. Bult, Jeffrey H. Chuang, Jill C. Rubinstein

**Affiliations:** The Jackson Laboratory for Genomic Medicine; Hartford Healthcare; Harvard School of Medicine; Department of Surgery, University of Pennsylvania; Yale School of Medicine; UConn School of Medicine; Johns Hopkins University; The Jackson Laboratory for Mammalian Genetics

## Abstract

**Purpose:** Pediatric sarcomas are rare and diverse, often leading to misclassification that hampers prognosis and treatment planning. We collected and harmonized histology images from multiple medical centers and developed accurate, generalizable classifiers for sarcoma subtype classification using deep learning techniques on digitized histology slides.

**Experimental Design:** A pediatric sarcoma histology dataset was amassed from three medical centers as well as the Children’s Oncology Group (COG), and a computational pipeline was implemented to address batch effects and standardize processing. Input parameter combinations (e.g. tile size, multi-scale feature extraction) were tested and optimized, and multiple convolutional neural network (CNN) and vision transformer-based (ViT) deep learning backbones were used as feature extractors for our SAMPLER whole slide image (WSI) representations, which we benchmarked to create an optimized, sarcoma-specific workflow. Models were trained for automated classification of rhabdomyosarcoma (RMS) versus non-rhabdomyosarcoma (NRSTS) and to further distinguish subtypes including alveolar, embryonal and spindle cell RMS as well as Ewing sarcoma.

**Results:** A total of 867 whole slide images were harmonized to generate a digital histology library with representation from 10 pediatric sarcoma subtypes. Across multiple classification tasks, ViT are superior to CNN models and multiscale feature sets consistently outperform single-scale models. Optimized classifiers accurately predicted RMS versus NRSTS (AUC 0.969±0.026) as well as alveolar versus embryonal RMS (AUC 0.961±0.021). Our two-stage classifier distinguished Ewing sarcoma from other NRSTS (AUC 0.929). Additionally, our models are computationally more lightweight than standard transformer implementations (model size 0.111 v. 1.9MB, training time > 3 orders of magnitude faster).

**Conclusions:** Digital histopathology can successfully classify pediatric sarcomas, providing results that are reproducible, mitigate inter-observer bias, and can be implemented remotely. We demonstrate that image harmonization overcomes the pitfall of overfitting to single institutional data while maintaining state-of-the-art performance across multiple classification tasks. The developed classifiers provide a basis for more precise histology-based sarcoma diagnostics, enabling global access to improved prognostication and treatment planning.

**Abstract:** Pediatric sarcomas present diagnostic challenges due to their rarity and diverse subtypes, often requiring specialized pathology expertise and costly genetic tests. To overcome these barriers, we developed a computational pipeline leveraging deep learning methods to accurately classify pediatric sarcoma subtypes from digitized histology slides. To ensure classifier generalizability and minimize center-specific artifacts, we collected and harmonized a dataset comprising 867 whole slide images (WSIs) from three medical centers and the Children’s Oncology Group (COG). Multiple convolutional neural network (CNN) and vision transformer (ViT) architectures were systematically evaluated as feature extractors for SAMPLER-based WSI representations, and input parameters such as tile size combinations and resolutions were tested and optimized. Our analysis showed that advanced ViT foundation models (UNI, CONCH) significantly outperformed earlier approaches, and incorporating multiscale features can enhance classification accuracy. Our optimized models achieved high performance, distinguishing rhabdomyosarcoma (RMS) from non-rhabdomyosarcoma (NRSTS) with an AUC of 0.969±0.026 and differentiating RMS subtypes (alveolar vs. embryonal) with an AUC of 0.961±0.021. Additionally, a two-stage pipeline effectively identified scarce Ewing sarcoma images from other NRSTS (AUC 0.929). Compared to conventional transformer encoder architectures used for WSI representations, our SAMPLER based classifiers were more lightweight (0.111 MB vs. 1.9 MB) and three orders of magnitude faster to train. This study highlights that digital histopathology paired with rigorous image harmonization provides a powerful solution for pediatric sarcoma classification. Our models reduce inter-observer variability, augment diagnostic precision, and have the potential to increase global accessibility to robust diagnostics, improving time to diagnosis and subsequent treatment planning.

**Statement of translational significance:** Digitized H&E-stained histopathology slides offer a transformative opportunity to develop artificial intelligence-based models for pediatric sarcoma histological classification. We leveraged advanced imaging analysis and deep learning techniques to develop classifiers capable of accurately identifying and differentiating subtypes of sarcoma. This innovation paves the way for imaging-based diagnostic methods that can be universally applied, ensuring that patients receive precise and timely diagnoses regardless of their geographic location or the resources available at their treatment centers. Consequently, this approach has the potential to significantly enhance the equity of access to precision diagnostics, ultimately improving prognostics and treatment planning for pediatric sarcoma patients worldwide.

## Introduction

Sarcomas are a clinically significant yet relatively rare group of mesenchymal cell cancers that present substantial challenges in diagnosis and classification. There are over 50 different subtypes of sarcoma, and their heterogeneity is amplified by the fact that they can arise in many different anatomical locations and by their varying biological behaviors^1,2^. Histopathologic evaluation of tumors using various staining and immunohistochemistry techniques is common, though genetic and molecular tests such as evaluation of fusion status often play a decisive role in classification of sarcomas into specific subtypes^3,4^. For example, in rhabdomyosarcoma (RMS), the most common pediatric soft tissue sarcoma, activating gene fusions involving *FOXO1* and either *PAX3* or *PAX7* result in aberrant transcription factors associated with a more aggressive clinical course. Genetic fusion status has largely replaced the historic histologic designation of alveolar subtype (as opposed to embryonal which is fusion negative) in clinical classification and staging protocols^5^. Nevertheless, these genetic and molecular tests can be costly, time-consuming, and access is often restricted to large medical centers with specialized personnel, thereby limiting their global availability. As a result, patients may experience disparities in care based on location or access to these resources^6,7^. It has been estimated that 25% of sarcoma diagnoses are discordant when reevaluated, illustrating the urgent need for improved diagnostic strategies for all patients^8^.

Recent advances in deep neural networks have shown promise for medical imaging tasks, including tissue segmentation, cancer classification, and prognostication^9,10,11^. These models have the potential to be used remotely and at scale, enabling clinicians and patients worldwide to access sophisticated diagnostic tools. However, robust performance of such models hinges on the availability of substantial, high-quality image datasets and requires careful attention to data curation, normalization, and architecture selection to avoid overfitting and achieve broad generalizability^12^. Prior machine learning approaches to sarcoma prediction have shown strong results, with AUROC values ranging from 0.85-0.95 for the task of alveolar vs embryonal classification^13,14,15^. However, these works have tended to focus either on RMS or non-RMS soft tissue sarcomas, but rarely both, and often have not included many of the rarer subtypes in their analyses. Therefore, increasing the size and heterogeneity of pediatric sarcoma datasets remains an unmet need both in the research field and for clinical applicability, where generalizability relies on sufficiently diverse datasets. Indeed Ho et al., showed that generalizing to external datasets is particularly difficult in sarcoma classification^16^.

To address these challenges, we have compiled a large and histologically diverse pediatric sarcoma dataset from St Jude Children’s, Massachusetts General Hospital, Yale New Haven Children’s Hospital, and the Children’s Oncology Group. The dataset spans ten different sarcoma types, and to our knowledge is the largest curated dataset of multi-institutional pediatric sarcoma images. This multi-institutional approach was specifically designed to minimize batch effects, staining inconsistencies, variations in microscopy and scanning methods, and other technical artifacts that could negatively impact model performance, while simultaneously enhancing representation across diverse subtypes^17,18^. To integrate the images from the various centers, we developed an efficient pipeline for processing hematoxylin and eosin (H&E) whole slide images (WSIs), incorporating key pathology practices such as multiscale viewing and artifact management.

We leveraged this standardized dataset to train deep learning models specifically designed for sarcoma classification. Our models make use of an optimized WSI representation approach (SAMPLER)^19^ for fast and accurate classification, providing a more globally feasible solution with lower storage demands and fewer training barriers. We test deep learning models as a function of varying image resolutions, tile sizes, and foundation models for feature extraction to assess for optimal parameters, and benchmark our method against the modern attention-based transformer architecture that has shown strong results in vision tasks including histopathology^20,21^. We envision that our simple, modular pipeline will pave the way for additional centers to contribute and analyze data on rare sarcoma subtypes, enabling further improvements to the classifiers and expanding the applicability of artificial intelligence in the diagnosis and classification of pediatric sarcomas.

## Results

### Dataset curation and attributes

We curated 5 separate datasets from three major institutions as well as the Children’s Oncology Group, containing 867 images of 10 different sarcoma subtypes from 722 patients, with associated metadata **Supplementary Table 1**. The dataset contains heterogeneity in the sarcoma subtype, anatomical location of the tumor, sex, and race **Supplementary Figure S1**. Additionally, the associated metadata included pathological diagnosis from the providing institute, which was used as ground truth for our classification tasks.

### Whole slide image processing

Each medical center uses unique tissue extraction and staining procedures, scanners, and image file formats. To unify the datasets from different institutes and reduce site specific batch effects we applied our established pipeline, validated in previous studies, which includes image format conversion, focus checking, resolution standardization, and stain normalization^22^. A schematic of the pipeline is shown in **Figure 1**, where we take images from each center, convert them to a standard image format, segment tissue areas of interest, perform data harmonization, divide the image into smaller tiles, and extract both deep learning and morphological features. Importantly, tiles are extracted at different sizes and resolutions, which we term here field of view (FOV), where the size and resolution are inversely proportional, and total pixels are fixed **Supplementary Figure S2A/C** and **Methods**. These tiles at different FOVs can then be used in various downstream applications. To demonstrate our method’s robustness, we clustered all of the tiles based on their features and show that the resulting clusters correspond to distinct regions of the original H&E image **Supplementary Figure S2B.** Similarly, we show how the morphological eccentricity values of different tiles, also extracted by our pipeline, vary across the original tissue **Supplementary Figure S2D**.

**Figure 1.**
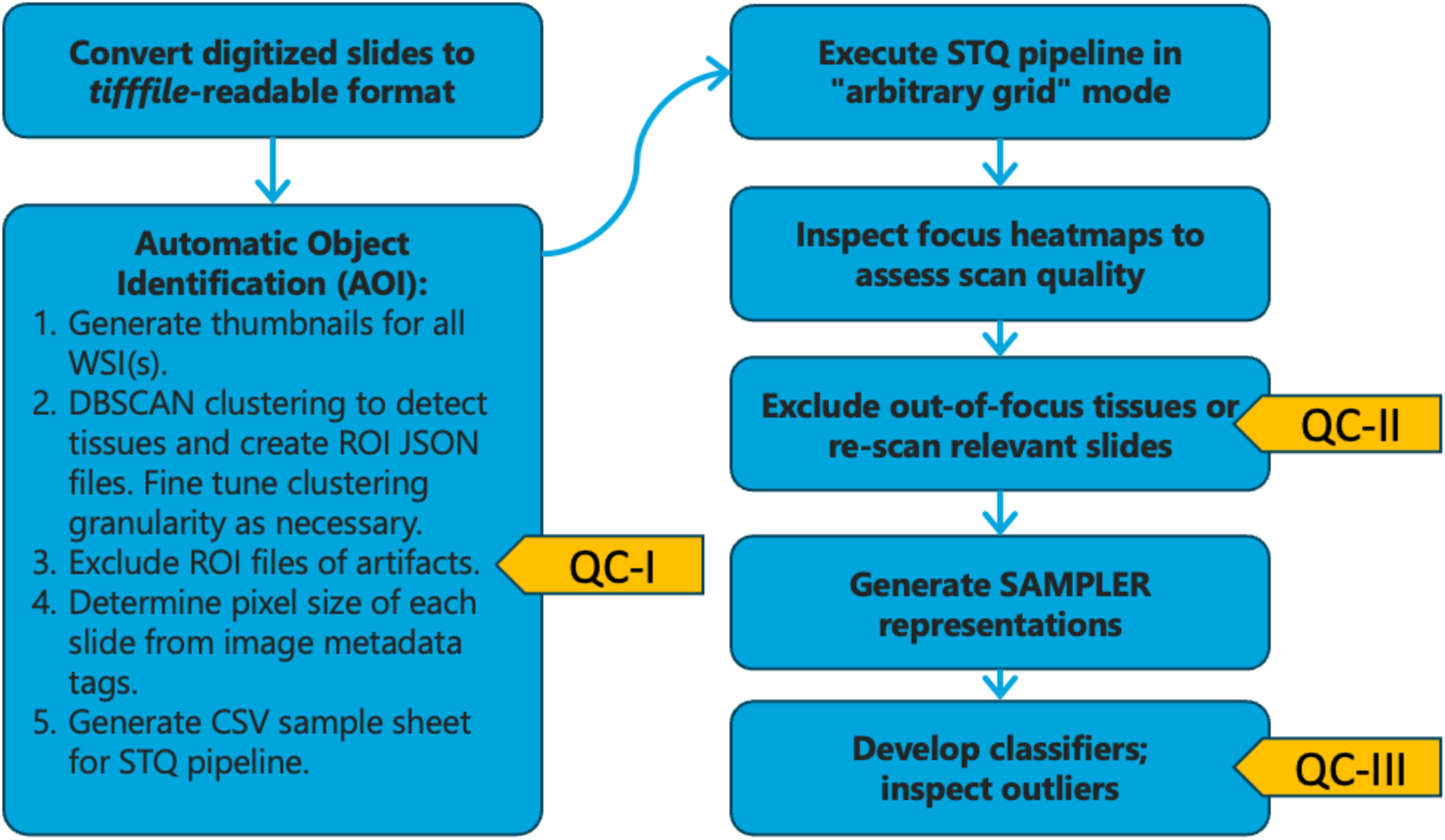
Workflow for processing whole slide images (WSI). The process begins with converting WSIs into a tifffile-readable format. The Automatic Object Identification (AOI) tool is then used to identify regions of interest (ROI) within the tissue samples, generating JSON files. Files of histology artifacts are filtered out. The STQ pipeline is executed in "arbitrary grid" mode to process each image and generate focus maps, tiling grid, imaging and nuclear morphometric features. The focus maps are examined for quality, with out-of-focus regions re-scanned or excluded. Finally, a SAMPLER WSI representation is generated for each tissue, followed by downstream classification analyses. Quality control has three distinct stages indicated with yellow pointers.

### Benchmarking of different feature extractors

Foundation models trained specifically on H&E-stained images have recently emerged as powerful tools for encoding histological features into abstract representations^23^. Because sarcoma subtyping often involves subtle morphological distinctions, we evaluated four such models as feature extractors to assess their potential for improving classification accuracy. We selected InceptionV3^24^, CTransPath^25^, UNI^26^, and CONCH^27^ to represent a range of established and newer architectures. Features from these four backbones were used to generate SAMPLER representations for downstream logistic regression. An important step in analyzing large whole-slide H&E-stained images is selecting an optimal tiling approach. To address this, we systematically examined tile size (a.k.a. patch size), image resolution, and tile overlap across multiple foundation models to determine optimal parameters for our classification tasks. Although higher-resolution images capture critical details such as nuclear mitoses, they can also introduce artifacts. In contrast, larger tile sizes provide a more extensive tissue context but may dilute finer morphological features.

To evaluate these parameters, we first tested them on the common task of alveolar versus embryonal RMS prediction, for which we also had the largest sample size. Alveolar RMS, associated with a poorer prognosis, was designated as the positive class because targeted therapies may be particularly effective against its fusion transcription factor. We found that the tile FOV had a substantial impact on classification performance across different deep learning backbones **Figure 2A**. For UNI, CTransPath, and InceptionV3, performance with the optimal FOV differed significantly from others. Notably, the smallest FOV (corresponding to the highest resolution) was not the top performer for most models. Additionally, the optimal FOV varied among backbones, reflecting differences in their feature spaces **Figure 2A**. Interestingly, for CONCH, a model trained on both image and language, differences in performance across fields of view were subtle and did not reach statistical significance **Supplementary Figure S3**.

**Figure 2.**
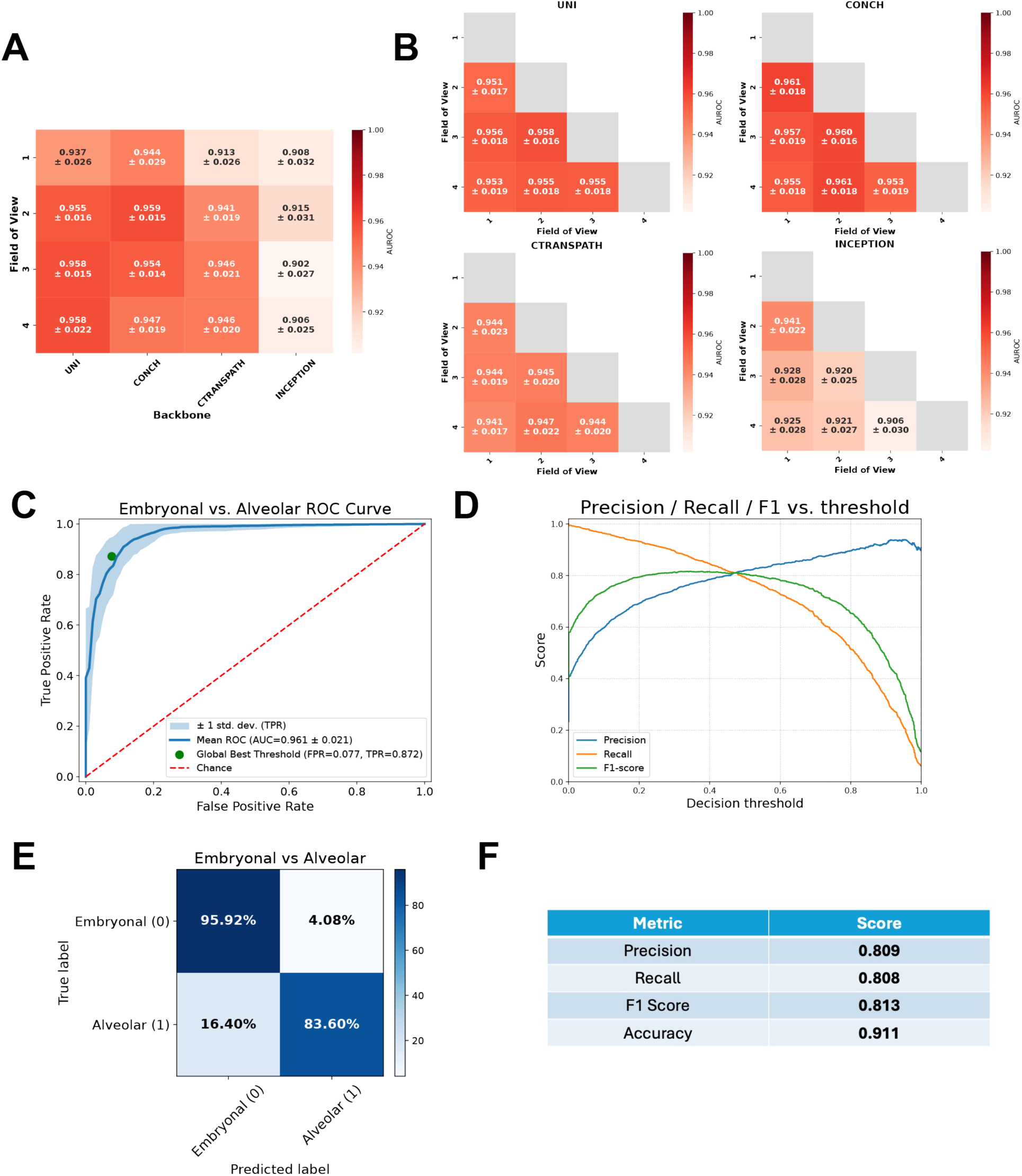
Benchmarking of deep learning backbones and image scales for pediatric alveolar vs embryonal sarcoma histological subtype classification. **A,** Mean area-under-ROC curve values for each of the 4 backbones at each FOV. **B,** Mean AUROCs from 5-fold cross validation with fixed random seeds for pairwise combinations of feature sets, where each feature set derives from a FOV for a given backbone. **C,** ROC curve for the best performing backbone combination, as measured over 100 5-fold cross-validation iterations. **D,** Plot of precision, recall, and F1 score curves for the best performing backbone combination (CONCH, FOV 1+2). **E,** Confusion matrix using the optimal threshold, averaged across all cross-validation iterations and expressed as percentages. **F,** Precision, recall, F1 score, and accuracy for the best performing backbone combination.

Overall, in the task of embryonal vs alveolar classification all four models achieved strong performance with our pipeline. ViT based models UNI and CONCH both produced state of the art results (AUROCs of 0.959 and 0.958, respectively), outperforming CTransPath (p < 5×10^-4^) and InceptionV3 (p < 5×10^-5^), with no significant difference between UNI and CONCH themselves **Supplementary Figure S4**. Notably, CTransPath also significantly outperformed the older convolutional neural network-based architecture InceptionV3. In the RMS versus NRSTS classification task, UNI and CONCH again surpassed the other two feature extractors (see also **Figure 4A**).

### Multiscale image resolution

Pathologists typically examine histology slides at varying magnifications to gain a more comprehensive understanding of tissue structures. Motivated by this practice, we investigated whether combining multiple scales for each deep learning backbone could enhance sarcoma subtype classification accuracy. Specifically, we concatenated the SAMPLER feature vectors extracted at two different fields of view, doubling the number of features used for downstream logistic regression. This multiscale approach yielded modest accuracy gains across all models **Figure 2B**. As an initial test for each FOV combination, we performed 5-fold cross validation using a set random seed to ensure robustness across comparisons. Notably, across all backbones and combinations, the vision-language model CONCH achieved the highest performance on alveolar versus embryonal classification using FOVs 1 and 2, corresponding to 40× resolution at 112 microns per pixel and 20× resolution at 224 microns per pixel, respectively. To further validate these results, we selected the best model parameters from the previous experiment and conducted 5-fold cross-validation over 100 iterations with varying seeds to ensure our models could learn with variable splits, which led to an improved state-of-the-art AUROC of 0.961 ± 0.021 **Figure 2C**. We report additional metrics for this model including precision, recall, and F1 scores across all thresholds **Figure 2D** and display the resulting confusion matrix, where we averaged the predictions across all test sets and express them as percentages **Figure 2E**. In clinical settings where inference is important the threshold used for final predictions can be adjusted to increase specificity for subtypes of interest. Importantly, despite the presence of class imbalance, the combined model maintained strong precision, recall, accuracy, and F1 scores **Figure 2F**. We also observed the model to be well calibrated, with predicted probabilities aligning well with actual predictions, which is important for potential clinical workflows **Supplementary Figure S5A**.

### Benchmarking of SAMPLER against transformer architecture

A growing trend in image classification is to use larger, deeper neural networks trained extensively on Graphical Processing Units (GPUs) to improve performance. Transformers encoders in particular offer a powerful way to generate WSI representations because they compute attention among all image tiles. Motivated by this, we assessed whether recently developed multi-head self-attention (MHSA) transformer architectures could outperform our SAMPLER-based classification approach.

To evaluate the strength of our trained models, we benchmarked the SAMPLER whole-slide image (WSI) representation and logistic regression model against a multi-head self-attention (MHSA) transformer architecture inspired by Wagner et al^21^. For each image tile, we extracted features using CTransPath and then employed a transformer encoder to calculate attention scores across all tiles. This strategy enables the model to capture both local and long-range spatial relationships, assigning hierarchical weighting to tiles based on their computed attention. In contrast, our standard SAMPLER approach summarizes tile-level features by calculating the empirical cumulative distribution function (CDF) of each feature and concatenating their quantile values to form a single vector. For this benchmarking experiment, features were extracted at a 20x magnification, and the transformer model was trained on an NVIDIA V100 GPU for 75 epochs, until training loss stabilized. To form the final WSI-level representation, global average pooling of the output attention scores was applied, yielding predictions as shown in **Figure 3A** and **Supplementary Figure S6**. Although transformer models can alternatively use class (CLS) tokens for aggregation in the final layer, both our findings and previous studies indicate that CLS tokens typically result in lower accuracy when training data is limited^28^.

**Figure 3.**
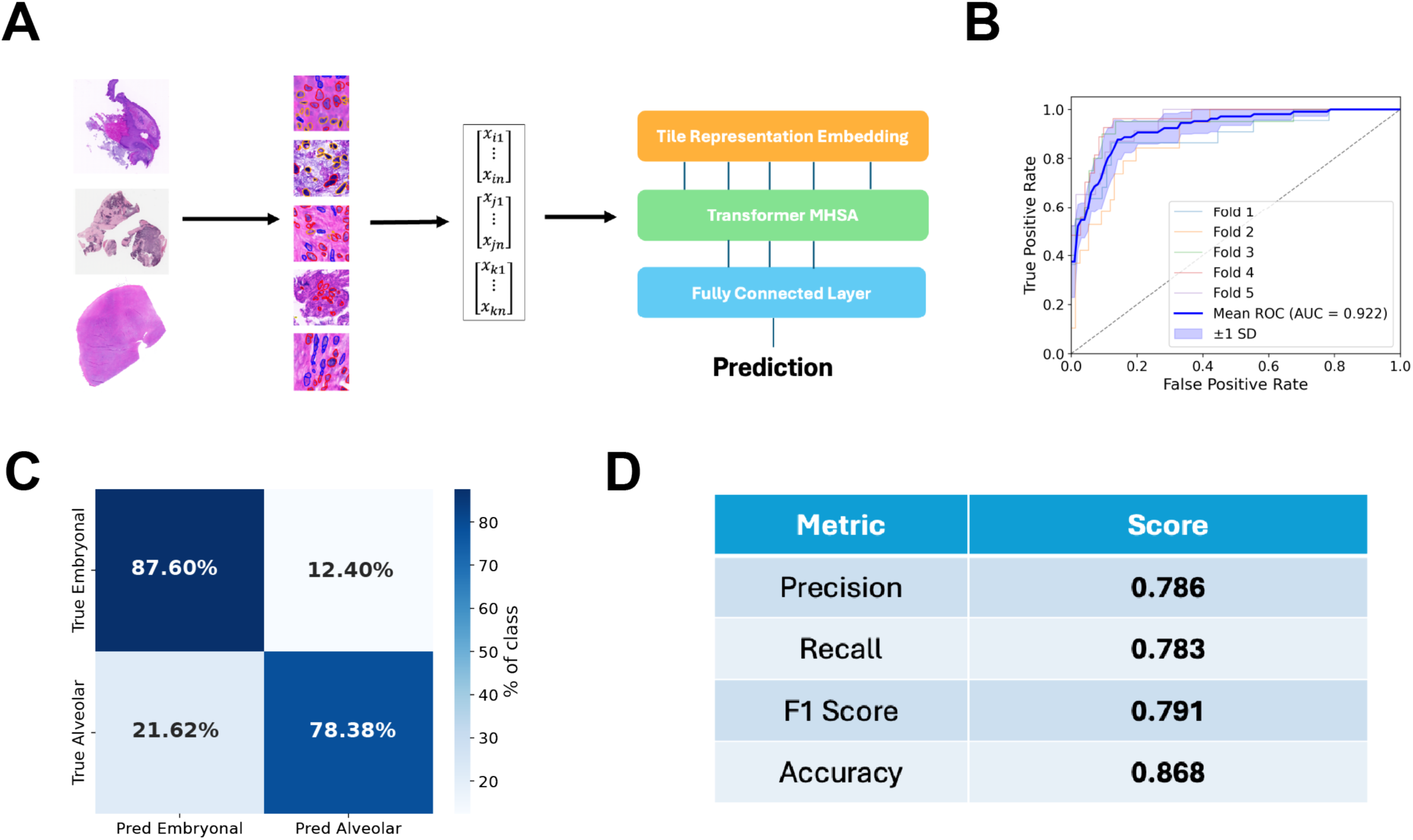
Alternative workflow using MHSA transformer architecture for pediatric sarcoma histological subtype classification. **A,** As a comparison to the SAMPLER and logistic-regression approach, we implemented the MHSA Transformer architecture to learn a WSI representation based on the interactions between tiles and then make classification predictions. **B,** ROC curve for the MHSA Transformer model in the task of Alveolar vs Embryonal classification. **C,** Averaged confusion matrix at threshold 0.5 for the Transformer. **D,** Precision, recall, F1 score, and accuracy metrics for the Transformer model.

In the alveolar versus embryonal RMS classification task, the transformer achieved a mean AUROC of 0.922 ± 0.031 across five folds **Figure 3B**. The average confusion matrix for each fold showed generally strong performance on both classes, though weaker than our SAMPLER models **Figure 3C**. Precision, recall, and F1 scores were relatively high, but also fell short of the SAMPLER-based method **Figure 3D**. When comparing model size, the transformer’s saved model (1.9 MB) was notably larger than SAMPLER’s (0.111 MB). Training time was also more than three orders of magnitude longer, even after setting aside five samples for inference testing **Supplementary Figure S7**. Because the transformer architecture underperformed SAMPLER and required substantially more computational resources, we proceeded with SAMPLER for subsequent tasks.

### RMS vs NRSTS histological subtype classification

Distinguishing RMS from NRSTS is a clinically important task that is not often addressed using machine learning due to the lack of datasets containing sufficient examples of the two distinct classes. Given the available heterogeneity in our dataset, we next attempted to train models for this task, which normally requires expert pathologist annotation in addition to a panel of molecular tests^29^. RMS, typically diagnosed by positive staining for protein markers such as myogenin and MyoD1, was designated as the positive class ^30^. Here, we used 66 NRSTS cases and 681 RMS cases, then trained across the entire dataset. We performed feature extraction using the various backbones at different tile sizes and resolutions and created SAMPLER representations as described above. We trained logistic regression classifiers on each combination of backbones and resolution scales, and compared the resulting 5-fold cross-validation averaged AUROC and accuracy values to determine the optimal parameters. The UNI foundation model with a FOV of 3 achieved the highest performance (AUROC 0.971 ± 0.022; **Figure 4A**). In the multiscale setting, the optimal combination of features was derived from the UNI backbone using FOVs 1 and 4 **Figure 4B**, though several FOV combinations showed overlapping performance within one standard deviation.

**Figure 4.**
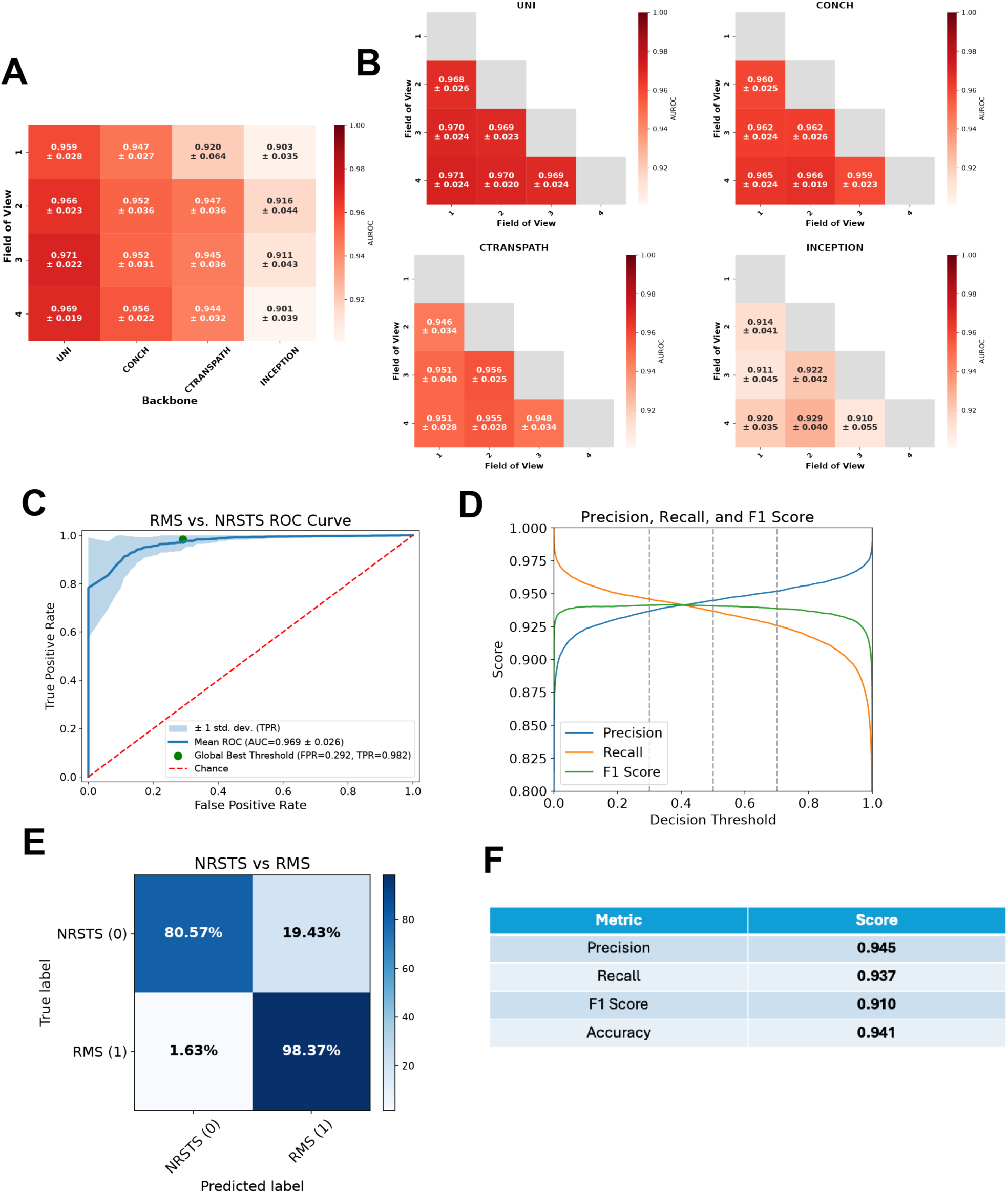
Benchmarking of deep learning backbones and image scales for pediatric RMS vs NRSTS sarcoma histological type classification. **A,** Comparison of mean AUCs for the 4 deep learning backbones at each FOV. **B,** Comparison of combinations of concatenated FOV features for each backbone, mean AUC. **C,** ROC curve for the best performing backbone combination over 100 iterations. **D,** Plot of precision, recall, and F1 score curves for the best performing backbone combination. **E,** Confusion matrix across all fold test sets, using the optimal threshold, expressed as percentages. **F,** Precision, recall, F1 score, and accuracy for the best performing backbone combination.

For further validation and to assess robustness across the dataset we performed 100 iterations of the 5-fold cross validation with the best backbone and combination pair (UNI 1+4). Across all iterations the model achieved an average AUC of 0.969 **Figure 4C**, and we additionally show the precision, recall and F1 scores across varying decision thresholds **Figure 4D**. We also report the confusion matrix, expressed in percentages, for predictions across all images in the dataset **Figure 4E**. Although RMS cases are predicted most accurately, likely due to their greater representation in the dataset, the model still effectively identifies NRSTS cases despite their relative scarcity. Additionally, we report the precision, recall, F1 score, and accuracy at the ideal threshold for accuracy across all folds **Figure 4F**.

### Classification of Ewing sarcoma histological subtype

Accurately recognizing minority classes in highly imbalanced datasets remains a major challenge for image-classification models, particularly in clinical settings where some pathologies are inherently rare. To evaluate our method under these conditions, we focused on Ewing sarcoma (ES), a tumor defined by the *EWSR::FLL1* fusion gene, which appeared in only 41 of the 722 cases in our cohort^31^. Multi-resolution features were extracted from the tiles of each image using several backbone networks and distilled into a single WSI SAMPLER vector as described earlier. Since ES falls under the NRSTS umbrella, we split the task into separate decisions to help account for the underrepresentation of ES cases.

To demonstrate how such a model might be applied clinically, we devised a two-stage pipeline to maximize interpretability **Figure 5A**. First, we trained a model with five-fold cross-validation to separate RMS from NRSTS across the entire dataset. All samples predicted as NRSTS were then fed into a second model designed to distinguish ES from other NRSTS subtypes. In the second stage, ES was chosen as the positive class, as targeted therapies are being developed for the fusion protein, making sensitivity for ES potentially more impactful^32^. We achieved high AUCs in both stages (0.959 and 0.938, respectively **Figure 5 C/D**), indicating that out approach could accurately identify ES despite the substantial class imbalance. To assess model performance realistically across both stages, we combined the predictions from each test set and summarized the results in a confusion matrix **Figure 5B**. Additionally, we report the precision and recall for both stages **Figure 5E/F,** as well as the overall metrics **Figure 5G**, underscoring the model’s effectiveness given the inherent difficulties of working with highly imbalanced datasets.

**Figure 5.**
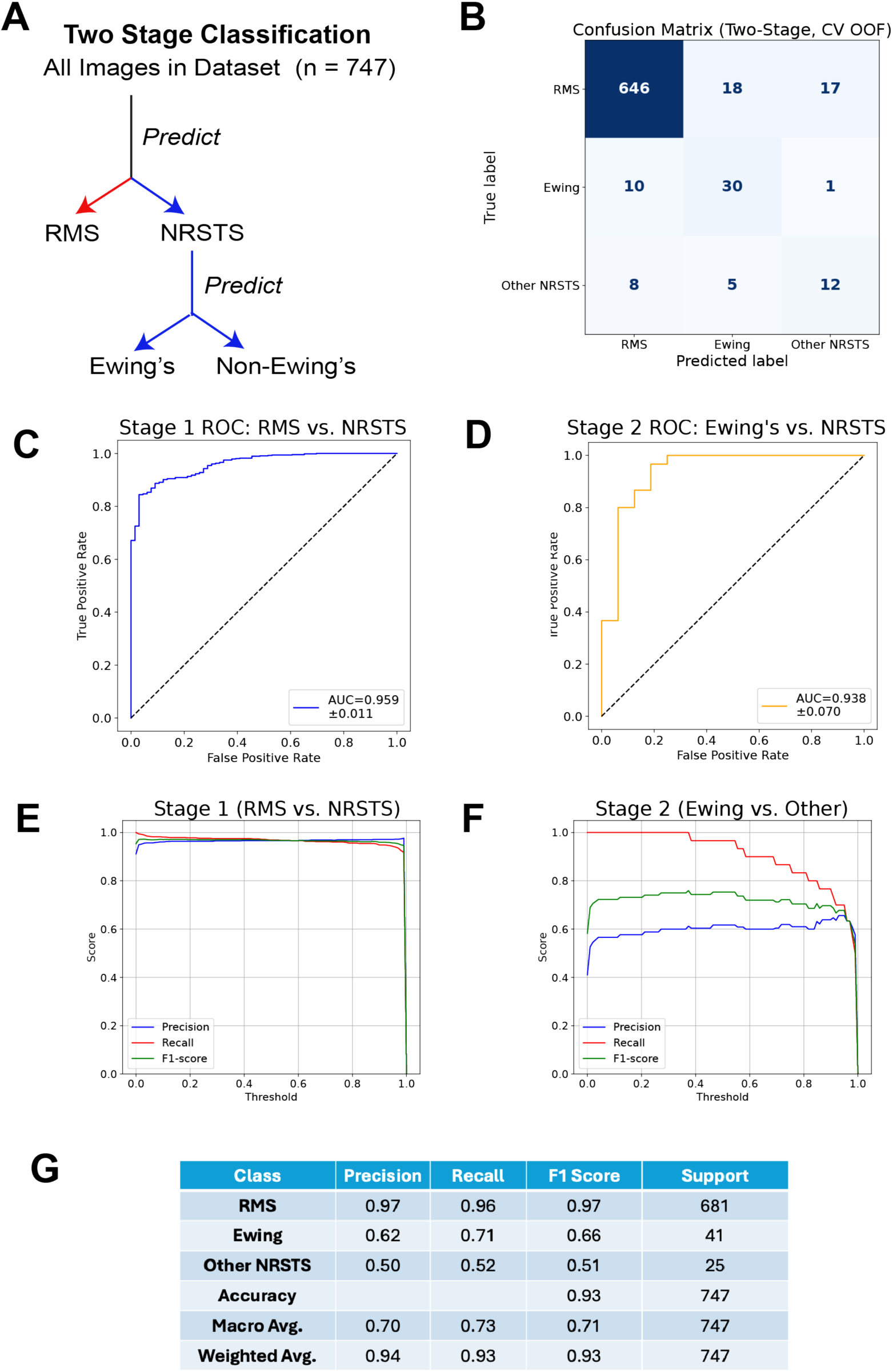
Two-stage classification of Ewing sarcoma. **A,** Schematic showing the two-stage classification workflow, where models are trained on all images in the dataset, and predicted NRSTS cases are moved to the second stage of classification. **B,** Confusion matrix for the combined stages is shown at threshold 0.8 (B). **C,** ROC curve for the first stage of classification NRSTS vs RMS. Any sample classified as NRSTS then moves to the next round for further subtyping. **D,** ROC curve for Ewing vs non-Ewing classification. **E/F/G,** Precision, recall, F1 score, accuracy and support for the two models.

### Multiclass classification of alveolar, embryonal, and spindle cell RMS

In applied clinical settings deep learning models need to be able to perform not only binary classification but extend predictions to multiple classes if necessary. We next tested our model’s ability to handle a multiclass prediction task that aimed to distinguish alveolar, embryonal, and spindle-cell RMS morphologies. Spindle tumors, while clinically and genetically distinct, are both rare and visually similar to the embryonal subtype, so they are seldom represented in RMS training sets^33^. As in the binary experiment, we extracted multi-resolution features with several backbone networks, collapsed each tile representation into a single SAMPLER vector, and trained logistic-regression classifiers in a one-versus-rest (OvR) scheme **Supplementary Figure S8A**. In this setup, three models are separately trained to identify their associated class, and the model with the highest predictive probability assigns the classification. This approach yielded a macro-average AUROC of 0.908, with class-specific AUROCs of 0.963 (alveolar), 0.893 (embryonal), and 0.869 (spindle) **Supplementary Figure S8B**. Because spindle morphology is more similar to embryonal then alveolar, the embryonal model is overly confident in calling spindle morphologies embryonal **Supplementary Figure S8C**. While this is technically correct, it is suboptimal for identifying the true spindle cases. To address this, we show that lowering the spindle decision threshold to 2.3 × 10⁻⁴ allows the model to uncover spindle-cell morphology within tumors otherwise labeled embryonal **Supplementary Figure S8D**.

### Spatial inference of histological types and subtypes

In addition to assessing classification performance, we sought to enhance the interpretability of our models by identifying specific morphological and structural features in H&E images that influence predictions. To accomplish this, we visualized tissue regions that most significantly impacted classifier decisions. However, identifying influential regions with the SAMPLER approach poses challenges, as SAMPLER disregards tile-level context and lacks a direct mechanism to quantify individual tile importance. Therefore, to analyze whether the model predictions were consistent with expectations from pathologist evaluations of H&E images, we developed a spatial inference approach based on local calculation of the SAMPLER representation **Supplementary Figure S9**. We then evaluated slides for local presence of sarcoma type and subtype signals using this approach. **Figure 6** illustrates the application of spatial inference on H&E-stained slides for differentiating between various subtypes of NRSTS and RMS images. Panels A–C illustrate classification tasks using examples of synovial NRSTS (A), alveolar RMS (B), and Ewing sarcoma NRSTS (C), respectively.

**Figure 6.**
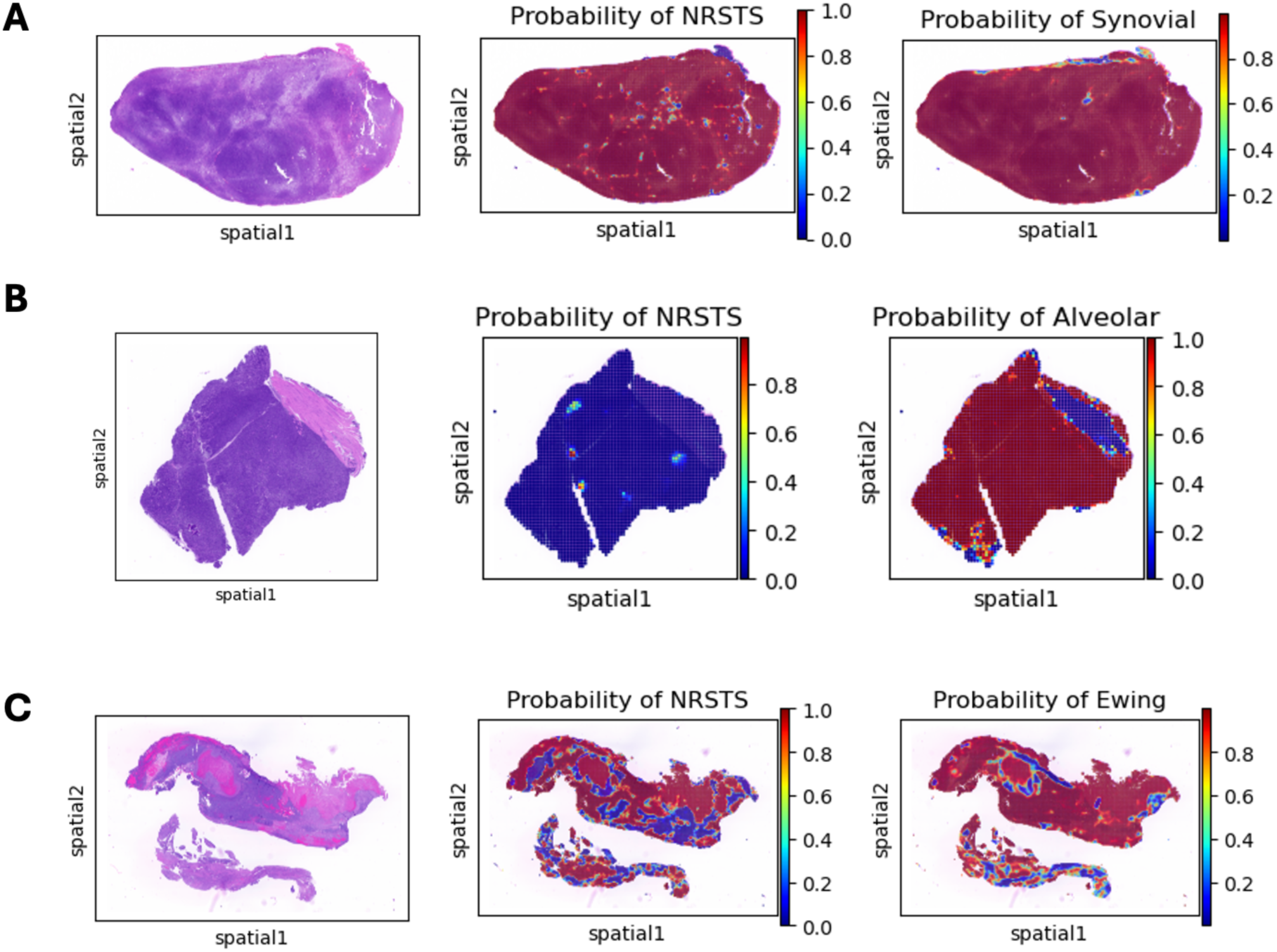
Spatial inference of histological types and subtypes of sarcoma. Panels A-C represent different subtypes of non-rhabdomyosarcoma soft tissue sarcomas (NRSTS) and rhabdomyosarcomas (RMS). Each panel includes three images: Hematoxylin and Eosin (H&E) stained tissue section (left), probability heatmap of NRSTS (middle), and probability heatmap of the specific subtype (right). **A,** Synovial NRSTS. **B,** Alveolar RMS. **C,** Ewing NRSTS. The heatmaps illustrate the spatial distribution of contributing features, which may aid in pathologist visual assessment and further validation by other assays.

The probability heatmap in **Panel A** indicates the likelihood of NRSTS in the middle image, and a probability heatmap for synovial sarcoma subtype is on the right. The tissue slide shows dominant composition of NRSTS and more specifically synovial sarcoma. **Panel B** shows that the classifier correctly assesses no part of the alveolar image as NRSTS and identifies the tumor region as being alveolar. The example in **Panel C**, diagnosed as ES NRSTS, exhibits certain RMS-like regions (highlighted in blue), which were confirmed through pathologist evaluation. Notably, all cases in Figure 6 align with the associated diagnostic ground truth, demonstrating the reliability of the approach.

## Discussion

For rare and diverse cancer subtypes encompassed by the sarcoma umbrella, establishing an accurate diagnosis typically requires extensive pathologic expertise alongside additional genetic and molecular testing. Deep learning approaches for analyzing H&E-stained images can help address this critical diagnostic challenge, potentially providing standardized accuracy and enhancing diagnostic capabilities in centers that lack the necessary expertise or resources to identify uncommon subtypes. Previous attempts to integrate digital image analysis into global clinical practice have been limited in part by computational resource constraints. To address this challenge we implemented a lightweight classifier designed for rapid training and inference, enabling it to run efficiently even on standard clinical computers. We validated our approach against a transformer architecture, which can have prohibitively large model sizes and training times^34^. While the accuracy of transformer-based models is competitive, they require the user to have expertise in training and running deep learning models due to the high optionality in hyperparameter design, as well as the potential need for GPU resources^35^. For this reason, other studies have explored how to reduce the memory and training requirements for transformers, while in contrast we applied a simplified approach for WSI representation that does not require computationally expensive attention calculations^36^. The SAMPLER method we employed addresses these concerns through a statistical transformation that approximates the features in an image as a single decile-based vector representation.

Calculating SAMPLER representations requires deciding which deep learning model to use as a feature extractor. This is important, as SAMPLER optimality may depend on the ability of the underlying features to encode the full range of visual biology within each tile. Indeed, the recent evolution of pathology feature extractors has led to greater accuracy in many classification tasks^23^. Here, we have profiled models including InceptionV3, a convolutional neural network trained on a general ImageNet dataset; a more recent H&E foundation model CTransPath; and one of the latest vision transformer models, UNI, trained on only H&E-stained images^24,25,26^. We verify that the evolution of feature extractors has led to gains in prediction accuracy in sarcoma cancer classification and further show that certain feature extractors are more suited for particular tasks, illustrating the importance of having a workflow that allows for flexibility in the backbone used. It is noteworthy but not surprising that CONCH, a model trained both on images and annotated clinical text that has excellent performance in other benchmarking tests^27^, is also the best in our alveolar vs embryonal task. This illustrates how creative architectures and differences in training may lead to continued improvements in the image foundation models. The STQ pipeline that is the core of our workflow allows for modular exploration of diverse feature backbones^22^. Our pipeline can be updated with other backbones as they become available, allowing groups to apply these models for specific tasks and finetune them on additional data.

In the training of our models, we aimed to mimic the approach of expert pathologists who view images through varying fields of view by zooming in and out across a slide. Previous groups have shown that a multiscale approach can provide more accurate models for segmenting images^37^. In addition to making the models more accurate, the multiscale approach might also make the models more robust to external validation by smoothing out certain artifacts when combining different views. Additionally, it is essential that computational researchers coordinate with pathologists to determine thresholds based on “can’t miss” diagnoses, to ensure clinically beneficial decision making^38^. For example, if a classifier is used to determine if a patient should receive a targeted therapy, the threshold should be set such that the specificity of the diagnosis being targeted is higher. On the other hand, certain subtypes are known to be less aggressive, and therefore classification of these subtypes should consider the balanced range of treatment options.

While deep learning approaches hold the potential to provide greater access to care and ease the healthcare burden globally, there are important caveats with their use. Many modern deep learning architectures are prone to overfitting, and this is especially true in pathology image analysis where there is a sparsity of data and each medical center could introduce specific artifacts^39^. These artifacts are usually ignored by pathologists, but deep learning models may internalize them, causing reduced generalizability. Our pipeline automates much of the image normalization between centers, while our curated sarcoma dataset contains diversity across institutions, studies, and patients, dampening the impact of artifacts and enabling multi-class predictions. There is still room for improvement, as the small number of images for the rarer subtypes limited our model’s ability to accurately classify them, as seen in the imbalanced class predictions (Ewing sarcoma, spindle cell RMS). More work is needed to expand this dataset and train additional models on these rarer subtypes. While we achieved state of the art results in multiple tasks, we also recognize that accuracies and AUROC values in part reflect the difficulty of the underlying dataset, therefore we look forward to testing and validating our models on additional images from external centers and collaborators.

## Conclusion

Our multi-institutional pediatric sarcoma histology dataset represents the largest harmonized resource of this type to our knowledge. We demonstrate the ability of histology foundation models to be integrated within a multiscale SAMPLER-based workflow to provide fast and accurate H&E image-based distinction between sarcoma subtypes. We achieve state of the art results for a number of clinically relevant tasks using a computationally efficient approach that could improve diagnostic accuracy and time to treatment. Additionally, our harmonization procedures open the door for expansion of the dataset through further multi-institutional collaborations, bringing promise for a future in which automated image review may accurately and remotely identify sarcoma histology, improving subtype-specific delivery of care.

## Methods

We collected Hematoxylin & Eosin (H&E)-stained images and limited clinical data from various treatment centers, including the Children’s Oncology Group (COG), St. Jude Children’s Research Hospital, Yale University, and Massachusetts General Hospital (MGH). We developed a pipeline to ensure focus checking, resolution standardization, stain normalization, and image format conversion, resulting in a harmonized dataset of 867 images (from 686 whole slides, 614 patients).

To optimize classification accuracy, we experimented with different tile sizes, overlaps, magnification powers, and single- vs. multi-scale concatenated-feature sets. Deep learning feature extraction was performed using five backbones: InceptionV3, CTransPath, UNI, and CONCH. We used our previously developed SAMPLER^19^ method to create statistical representations of each feature, which were then used to train and test artificial neural network (ANN) classifiers for distinguishing between RMS and NRSTS (Non-Rhabdomyosarcoma Soft Tissue Sarcomas), as well as further subtype predictions. For the SAMPLER analyses, we used decile statistics (5th–95th percentiles) to parameterize the feature distributions within tiles of each WSI. We compared the performance of these four backbones and benchmarked the resulting SAMPLER representations of features using Logistic Regression, Support Vactor Machine classifiers, and a transformer-based deep learning classifier.

### Slides processing workflow

The processing of whole slide images (WSI) is accomplished in batches involving several methodical steps. Initially, the WSIs are converted into a format readable by the tifffile Python library^40^, if WSIs are not already readable by the library. Following this conversion, we use our Automatic Object Identification (AOI) tool to identify regions of interest (ROI) within the tissue samples, generating corresponding JSON files. The tool is supplied and maintained within the Spatial Transcriptomics Quantification (STQ) pipeline^22^ repository, developed by The Jackson Laboratory. The raw ROI JSON files are then scrutinized to filter out any unnecessary tissue regions, imaging or histology artifacts, refining the dataset to include only relevant tissue regions. Subsequently, the STQ pipeline is executed in the “arbitrary grid” mode on the refined set of tissues to perform further analysis. Automation is a key aspect, once launched, STQ streamlines parallel processing of tissue images and eliminates any manual intervention. Within the pipeline run, the focus heatmaps are generated by the microscope scanner for each tissue according to Deep Focus^41^ with a custom-tuned checkpoint and are then examined to assess the quality of the scans. If any tissue regions are found to be out-of-focus, the slides are re-generated following our WSI scanning protocol. In cases where re-scanning is not feasible, the affected slides are excluded from downstream analysis to maintain data integrity. Finally, a SAMPLER representation^19^ with 10 quantiles ranging from 0.05 to 0.95 is generated for each tissue, providing a concise representation of the tissue samples for downstream classification analyses.

### Quality control

Our WSI data analysis includes three stages of quality control (QC). During stage I we exclude from further processing any WSI regions that correspond to histology artifacts, such as tissue folds or other microtomy artifacts, mounting media blobs, excessive contamination and air bubbles. During stage II, when STQ processing is complete, we exclude any slides with out-of-focus tissue regions given that re-scanning of such slides is infeasible. During stage III, we identify any outliers stemming from the machine learning analysis and investigate the clinical metadata for such cases.

### Automatic Object Identification

The Automatic Object Identification (AOI) workflow begins with the generation of thumbnail images for all whole slide images (WSI), providing a quick visual overview of the dataset as well as input to the next step. Next, the DBSCAN clustering algorithm is employed to identify tissues and their replicates within the WSIs, creating of regions of interest (ROI) JSON files. During this step, the granularity of the clustering may be adjusted as necessary to ensure accurate identification of tissue regions, tissue replicates and background padding around the tissues. Any ROI files that correspond to imaging or histology artifacts, such as tissue folds, mounting media blobs and air bubbles, are excluded from further analysis. This is stage I of quality control (QC) in WSI data analysis. The next step involves exploring the image metadata to determine the pixel size of each slide. Pixel size refers to the physical dimensions of a single pixel in the image, measured in micrometers (µm), and it determines the resolution of the image. Finally, a CSV sample sheet is generated for input into the Spatial Transcriptomics Quantification (STQ) pipeline, ensuring that all necessary metadata and ROI information are organized and ready for subsequent processing.

### Tissue region of interest definition file

The JSON file defines regions of interest (ROI) within an image by specifying two key attributes for each region: location and size. The location attribute indicates the position of the corner of the ROI within the WSI, using normalized coordinates that range from 0 to 1, representing a relative position along the image’s dimensions. The size attribute describes the relative size of the ROI, also using normalized values from 0 to 1, which represent the proportion of the image area that the ROI occupies. Together, these attributes allow for precise identification of a specific region within the image for further analysis.

### Pipeline implementation and architecture

In this work we used Spatial Transcriptomics Quantification (STQ) pipeline v3.0.0^22^. The improved architecture of this version of STQ, from a software engineering perspective, is optimized to be multi-functional, modular, scalable, and efficient, facilitating the processing of H&E-stained WSIs. Additionally, STQ v3.0.0 includes utility Automatic Object Identification (AOI) for preparation of the STQ input sample sheets.

The new pipeline version features a modular Nextflow design with distinct components for tasks such as region of interest (ROI) extraction, image conversion, image focus quantification for quality control, and data representation in a standardized output structure. The pipeline includes several deep learning (DL) models, and the pipeline architecture is extensible, allowing for the integration of new DL models and tools to meet the evolving needs of spatial biology research.

### Tile size and scale

Similar to the SAMPLER work^19^, we implemented multi-scale imaging feature extraction as follows. An image tile of the desired size is read from the tissue image and resized to the deep learning model’s required size using the "resize" transformation from TorchVision. The base tile size is defined by the parameter "grid_spot_diameter." The pipeline accepts a set of input scales, defined as a list of integers (e.g., 1, 2, 3, 4), which determine the size of the tile to read in multiples of the base size. This approach allows profiling different tile scales centered at the same coordinate but spanning varying vicinities. Note that nuclear segmentation results are aggregated only for the base tile size. Tile overlap is implicitly defined by the base tile size, center-to-center spacing, and grid shape, which can be square, hexagonal, or random.

### Feature sets extracted with the pipeline

For each tissue, we generate four imaging feature sets and a nuclear morphometric feature set. The imaging feature sets were generated from the following DL models: InceptionV3^24^, a convolutional neural network (CNN) architecture from the Inception family, was pretrained on ImageNet by Google to efficiently handle complex image recognition challenges. CTransPath^25^ is a transformer-based model for histopathological image classification, pretrained using unsupervised contrastive learning on histopathology images. UNI^26^ is a general-purpose self-supervised model for computational pathology, pretrained on over 100 million images from diverse tissue types to enhance performance across various clinical tasks. CONCH^42^ is a vision-language foundation model for histopathology, pretrained on a large dataset of image-caption pairs to support tasks like image classification, segmentation, and retrieval. These models are designed to leverage large-scale data and advanced architectures to improve the accuracy and efficiency of histopathological analysis. All these networks have implementations in PyTorch, and some, e.g., InceptionV3 and UNI have also TensorFlow implementation. We used these frameworks to include them in the STQ pipeline for image analysis.

### Tissue image classification using SAMPLER representation

Extracted features are processed as described in the SAMPLER paper^19^. In brief, for each extracted feature the empirical cumulative distribution function (c.d.f.) is assessed across all tiles of the image and is represented as 10 quantiles (5^th^…95^th^). The quantile representations of the features are concatenated together as a single vector, therefore ignoring tile level context. Downstream, we use and test both Logistic Regression (LR) and a Linear Kernel C-Support Vector of Support Vector Machine (SVM) to train the classifiers on the sets of image vectors. LR coefficients are easy to interpret, while SVM is effective in high-dimensional spaces and is robust to outliers, controlled by the value C for the optimization of the support vectors.

To ensure robust and unbiased evaluation of our model, we stratify the train-test datasets using abstracted patient identifiers. This stratification helps maintain the distribution of patient characteristics across both datasets, preventing any potential data leakage and ensuring that the model’s performance is not artificially inflated. For initial comparisons we then employ a five-fold Monte Carlo cross-validation procedure, which involves repeatedly splitting the data into training and testing sets (80%/20%) to estimate the mean and standard deviation of the classification performance. For the best performing models, we further validate by repeating 5-fold cross validation 100 times with varying splits to ensure robustness. The performance of our binary classification tasks is quantified using the Area Under the Receiver Operating Characteristic curve (AUROC), a metric that evaluates the model’s ability to distinguish between the two classes. The desired AUROC scores range from 0.5 to 1.0, with a higher score indicating better classification performance, while a score below 0.5 suggests that the classifier is performing worse than random guessing. We also measure accuracy, precision, recall, and F1 score to better estimate

### Transformer based image classification

We modeled our transformer encoder architecture similarly to Wagner et al^21^ with a few modifications. In brief we trained a multi-head self-attention transformer-encoder model on the extracted features of the tile sets for each image. Each slide was split into 224×224 tiles at 20x (0.5mpp) and each tile was fed into the CTransPath deep learning backbone which represents each tile as 768-dimensional feature vector, extracted from the last hidden layer. Each tile vector is then stacked together, and each individual slide is represented as *X* ∈ *R*^*n*×𝟟𝟞𝟠^, with n representing the number of tiles for that slide, which is variable. Each tile is then projected to a lower dimensional space of 512 through *z* = *ReLU*(*XW*_*proj*_ + *b*_*proj*_), *W*_*proj*_ ∈ ℝ^(768×512)^. To incorporate spatial context into the transformer model, we added a positional embedding to the original tile feature vectors. Specifically, for a given dimension *d* in the embedding space, positional values were computed as:

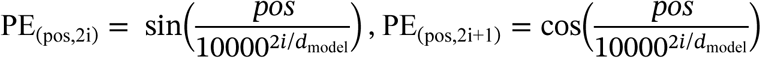

where *pos* represents the tile index, and *i* is the embedding dimension index. During each forward pass the positional encoding for the current sequence length is added to the learned tile embeddings, enabling the model to capture spatial order. Every tile vector (768 dimensions) is then linearly projected, followed by a ReLU, down to 512 dimensions, yielding an *n x* 512 sequence. This sequence is processed by three Transformer encoder layers, each with four heads and a position-wise feed-forward network with hidden width 1024. Within each layer, a multi-head self-attention block is followed by residual addition, layer normalization, and dropout (0.1) to mitigate over-fitting. Self-attention relates every tile to every other tile via a softmax calculation:

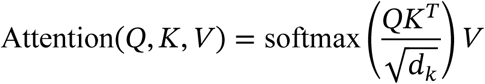

Queries *Q*, keys *K*, and values *V* are computed as *Q* = *W*_*Q*_ *x*, *K* = *W*_*K*_ *x*, *V* = *W*_*V*_ *x*, with *W* being a learnable variable optimized during training.

After the final layer, a fully connected layer with global pooling was used for the output prediction. Additionally, we tested whether appending a randomly initialized CLS token would improve accuracies, with similar performance. The model was trained using a binary cross-entropy loss function with logits (BCEWithLogitsLoss) weighted to compensate for class imbalance, calculated based on the ratio between the majority and minority classes.

All transformer-based experiments were performed using a NVIDIA V100 GPU (32G). The Adam optimizer was employed with a learning rate of 1e-5. Training was done 75 epochs per fold (as training loss began to plateau). Data was split into training and validation sets using stratified group 5-fold cross-validation to ensure balanced representation across classes and prevent patient data leakage between splits.

### Spatially resolved class probability heatmaps

In a manner similar to the WSI SAMPLER representation, we construct a local SAMPLER representation for each image tile. This involves selecting the tile and its nearest neighbors in the spatial coordinates within a specified radius, R. For instance, with R=2, we include the tile and its 12 surrounding neighbors to create an empirical c.d.f. with 10 quantiles ranging from 0.05 to 0.95 for each imaging feature. This local SAMPLER representation is then used for inference by employing the classifier trained on the WSI SAMPLER representation. Notably, increasing the radius results in smoother maps, as it incorporates more neighboring tiles into the analysis. With a setting of R=0 only one tile is used in constructing a c.d.f., resulting in a step function where each quantile of a given feature has the value of that tile feature.

Using spatially resolved class probability heatmaps, scrutinizing spatially distant regions of a WSI separately is beneficial because it allows for the identification of tissue heterogeneity, enabling pathologists to understand varying characteristics within the sample. This focused analysis can highlight critical areas, such as tumor margins or regions of inflammation, leading to more precise and accurate diagnoses. Additionally, breaking down the WSI into smaller sections enables the integration of other data types, such as spatial transcriptomics and spatial antibody-based imaging. Such data are often available only for tissue regions smaller than a full H&E-stained WSI.

### Two-dimensional view of the slide collection

We utilize the SAMPLER representation of all imaging features to compute the top 100 principal components with the largest variance, using the Python package Scanpy. These principal components are then zero-centered to standardize the data. To correct for batch effects, we apply the Harmony algorithm,^43^ using the four centers as the batch variable. This ensures that variations due to different centers are minimized. After batch correction, we compute the 8 nearest neighbors for each data point using the correlation similarity metric. Next, we generate a Uniform Manifold Approximation and Projection (UMAP) to visualize the data in a lower-dimensional space. This UMAP helps in exploring the clinical metadata overlaid on the visualization, clarifying how different clinical variables relate to the imaging features.

### Histology whole slide images scanning guidelines

To assist with histology image quality control, we use the following guidelines. When scanning whole slide images, ensure the resolution is approximately 0.25 ± 0.05 microns per pixel (mpp) to capture detailed cellular structures. Make sure the scan region spans the entire slide including the tissue background. With the default settings, the scanner may save slide “thumbnail” image and label image. Ensure that the label image does not contain any protected health information (PHI); remove the label image from the digital slide if PHI is captured.

Calibrate the scanner before use to ensure accurate focus and use automatic or manual focus adjustments as needed to maintain clarity across the entire slide. Check for uniform illumination and color balance, and ensure there are no artifacts, e.g., vertical or horizontal bands, grid patterns, or distortions in the scanned image. Additionally, ensure the slide is clean and free of any dust or debris, properly mounted, and cover-slipped.

When saving the images use either the recommended scanner compression setting or set it to use lossless compression to maintain image quality. Scanned images can be saved in a high-quality file format specific to the vendor, e.g., Aperio ScanScope Virtual Slide or SVS for Aperio by Leica Biosystems, NDPI for Hamamatsu NanoZoomer, etc. Using available software, e.g., ImageScope by Leica Biosystems, review scanned images for consistency and accuracy, verify that in the maximum magnification the image retains its detail and clarity.

### Abstracted clinical data acquisition protocol

To assist with clinical data quality control, we use the following guidelines. When submitting whole slide images (WSI), please ensure that the following metadata is provided: abstracted patient ID, abstracted slide ID, histological type, histological subtype, anatomical location, patient age at the time of sample collection, patient sex, patient race and ethnicity, primary tumor or metastatic site status, and a molecular signature summary. If available, include a pathologic description or summary of findings concerning the histology slide. This comprehensive metadata will facilitate accurate analysis and interpretation of the WSIs, ensuring that all relevant clinical and pathological information is available for use with digital pathology machine learning algorithms. The metadata can be submitted in either an excel table or plain text format. Please ensure that no Protected Health Information (PHI) is included to maintain patient confidentiality and comply with privacy regulations.

### Software availability

The STQ pipeline v3.0.0 is freely available at https://github.com/TheJacksonLaboratory/STQ. SAMPLER is freely available at https://github.com/TheJacksonLaboratory/SAMPLER

## Data Availability

All data produced in the present study are available upon reasonable request to the authors.

## Acknowledgements

This project was supported by a supplement to NCI Cancer Center Support Grant P30CA034196 as well as NCI grants U24 CA224067, U24CA263963, and R01CA230031. We acknowledge the use of artificial intelligence services ChatGPT to revise and edit the text document. These services were not used to generate novel text. Further we acknowledge the use of Github Copilot and ChatGPT to both write and edit parts of the code used in this manuscript, which was checked and edited by the authors.

## Supplementary Tables and Figures

**Supplementary Figure S1.**
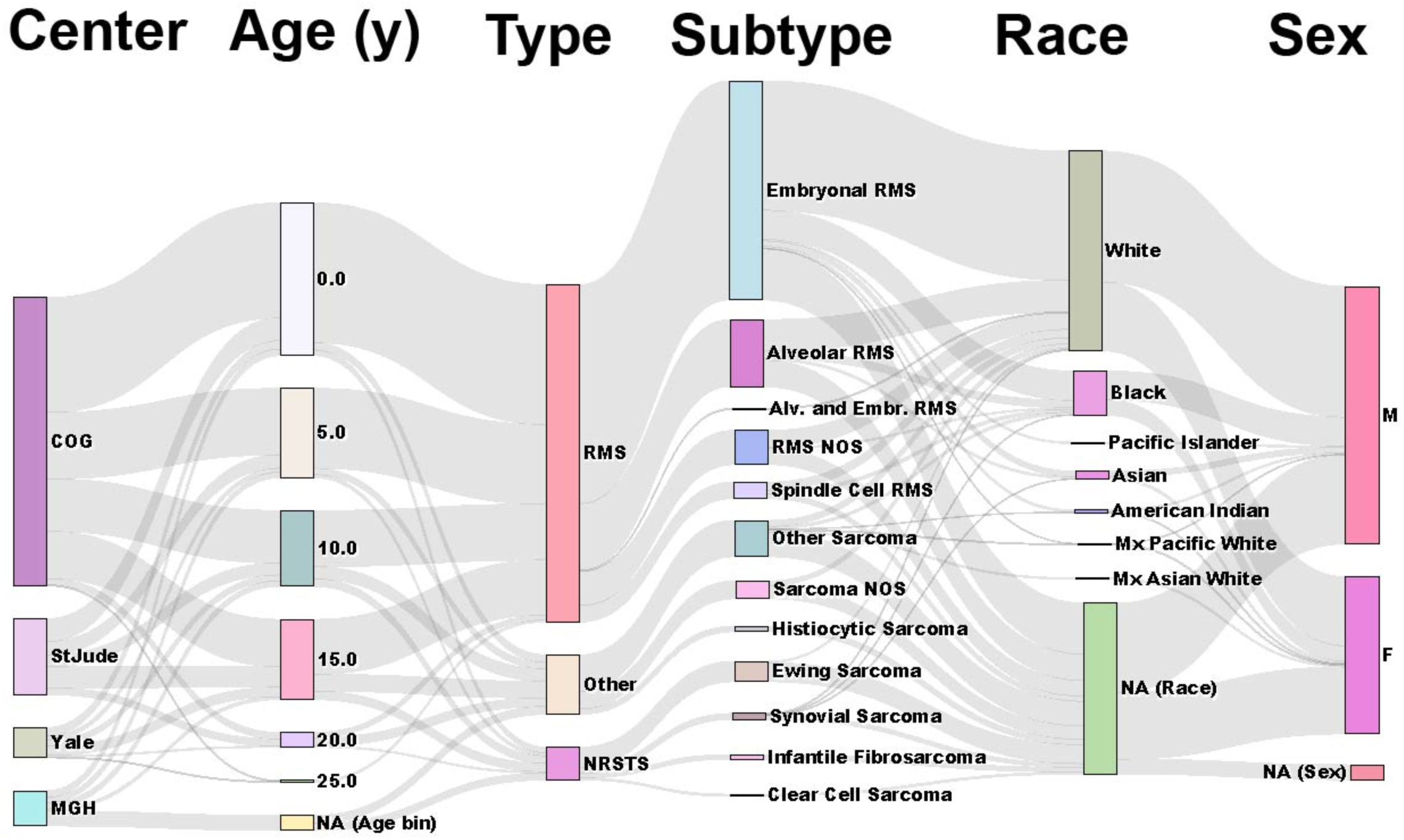
Metadata connection across institutes. Schematic showing the diversity of the curated dataset from left to right: institution where image originated, age bin (years), broad histological type, specific histological subtype, race, and sex.

**Supplementary Table 1.** Counts of cases, slides, and images are shown for each histological subtype included in the dataset. STJ, St. Jude Children’s.

|  |  | Patients |  |  |  |  | Slides |  |  |  |  | Images |  |  |  |  |
| --- | --- | --- | --- | --- | --- | --- | --- | --- | --- | --- | --- | --- | --- | --- | --- | --- |
| Histological Type | Histological Subtype | COG | MGH | STJ | Yale | Total | COG | MGH | STJ | Yale | Total | COG | MGH | STJ | Yale | Total |
| NRSTS | Clear Cell Sarcoma | 1 | 0 | 1 | 1 | 3 | 1 | 0 | 1 | 2 | 4 | 1 | 0 | 1 | 2 | 4 |
|  | Ewing Sarcoma | 3 | 5 | 0 | 13 | 21 | 3 | 11 | 0 | 19 | 33 | 3 | 18 | 0 | 20 | 41 |
|  | Infantile Fibrosarcoma | 0 | 1 | 1 | 3 | 5 | 0 | 1 | 1 | 5 | 7 | 0 | 1 | 3 | 5 | 9 |
|  | Synovial Sarcoma | 4 | 1 | 2 | 1 | 8 | 4 | 1 | 4 | 1 | 10 | 4 | 1 | 6 | 1 | 12 |
| Other | Histiocytic Sarcoma | 0 | 1 | 0 | 0 | 1 | 0 | 4 | 0 | 0 | 4 | 0 | 11 | 0 | 0 | 11 |
|  | Other Sarcoma | 20 | 1 | 17 | 2 | 40 | 20 | 1 | 27 | 3 | 51 | 29 | 1 | 40 | 3 | 73 |
|  | Sarcoma NOS | 7 | 4 | 0 | 6 | 17 | 7 | 8 | 0 | 9 | 24 | 12 | 14 | 0 | 10 | 36 |
| RMS | Alv. and Embr. RMS | 2 | 0 | 0 | 0 | 2 | 2 | 0 | 0 | 0 | 2 | 2 | 0 | 0 | 0 | 2 |
|  | Alveolar RMS | 84 | 0 | 13 | 8 | 105 | 84 | 0 | 18 | 10 | 112 | 91 | 0 | 33 | 11 | 135 |
|  | Embryonal RMS | 321 | 0 | 20 | 3 | 344 | 321 | 0 | 39 | 4 | 364 | 371 | 0 | 66 | 5 | 442 |
|  | RMS NOS | 32 | 7 | 0 | 3 | 42 | 32 | 11 | 1 | 4 | 48 | 39 | 25 | 1 | 4 | 69 |
|  | Spindle Cell RMS | 25 | 0 | 1 | 0 | 26 | 25 | 0 | 2 | 0 | 27 | 30 | 0 | 3 | 0 | 33 |
|  | Total | 499 | 20 | 55 | 40 | 614 | 499 | 37 | 93 | 57 | 686 | 582 | 71 | 153 | 61 | 867 |

**Supplementary Figure S2.**
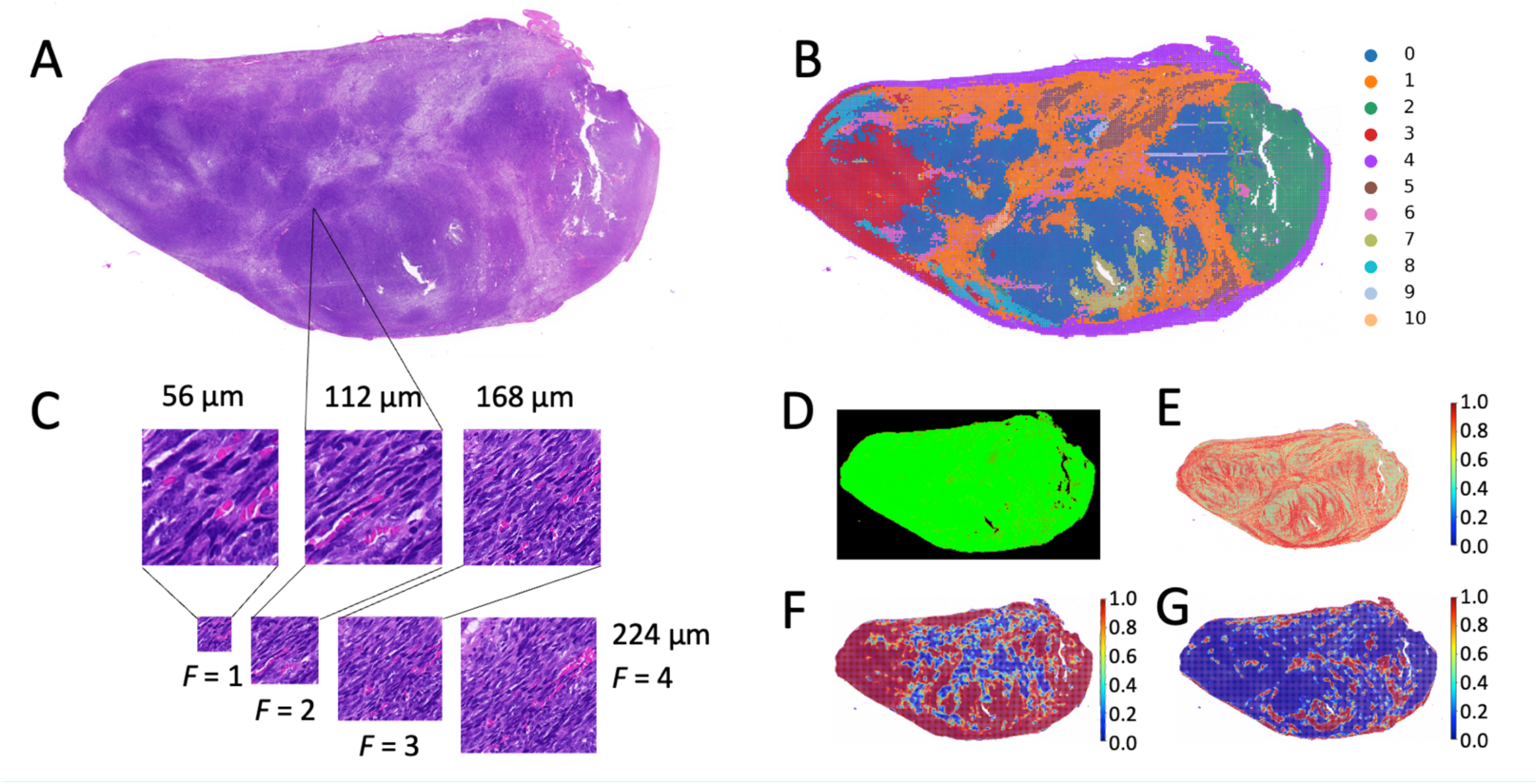
Case of synovial sarcoma chest metastasis in a pediatric patient. **A,** Histology H&E-stained WSI. **B,** Clustering-based segmentation of CTransPath features, overlaid on the original H&E image. **C,** Examples of tiles of varying sizes and resolutions, illustrating field of view (FOV) differences. **D,** Focus map: green indicates good focus, red indicates poor focus, and black indicates non-tissue areas. **E,** Average eccentricity of nuclei per tile at scale factor F = 1 (FOV 1). **F,** Regional NRSTS class probability at radius R = 2 (see methods). **G,** Regional RMS class probability at radius R = 2.

**Supplementary Figure S3:**
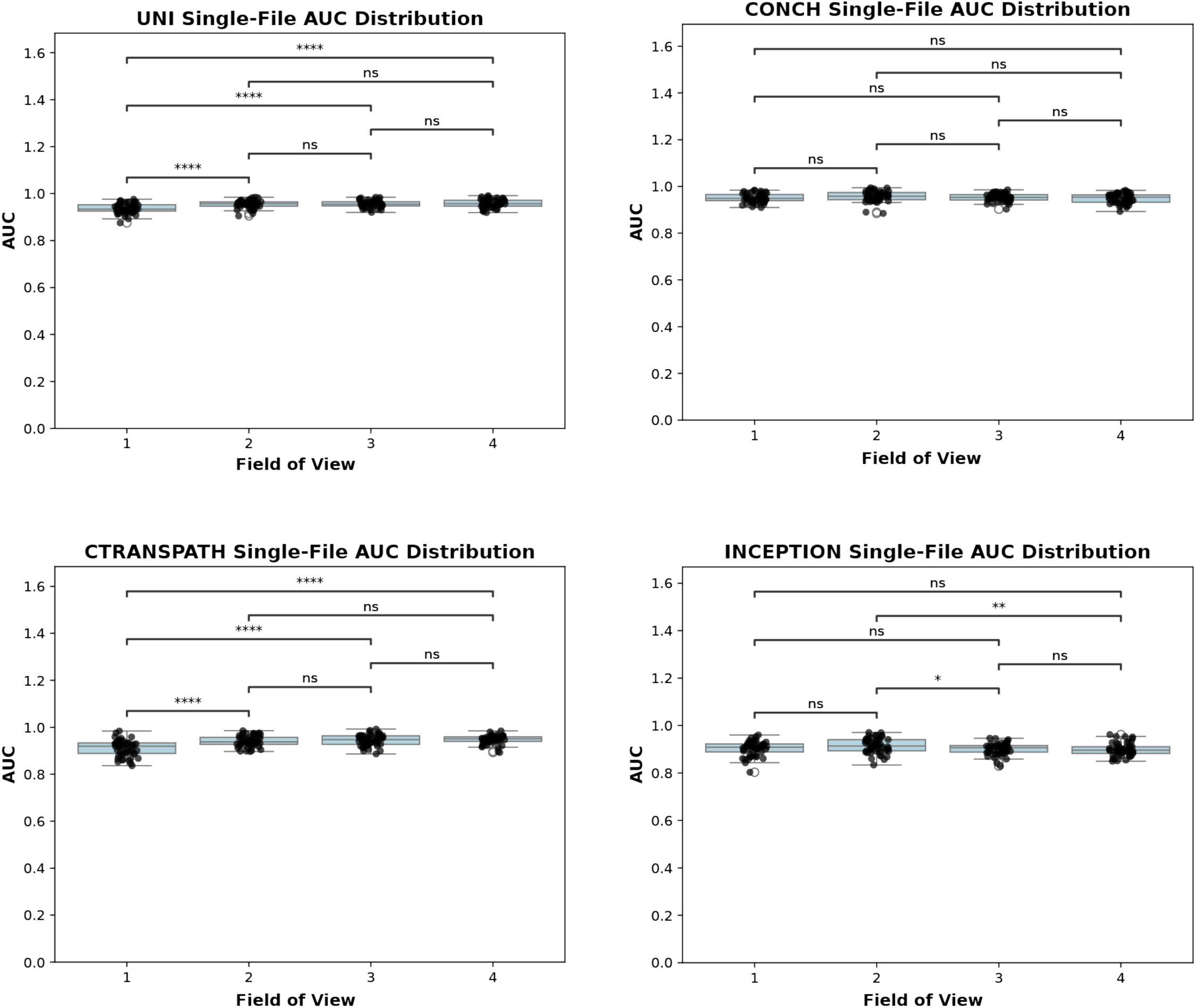
Differences in each backbone’s performance in the task of embryonal vs alveolar classification at different FOVs. 5-fold cross validation was repeated 50 times with varying random seeds.

**Supplementary Figure S4:**
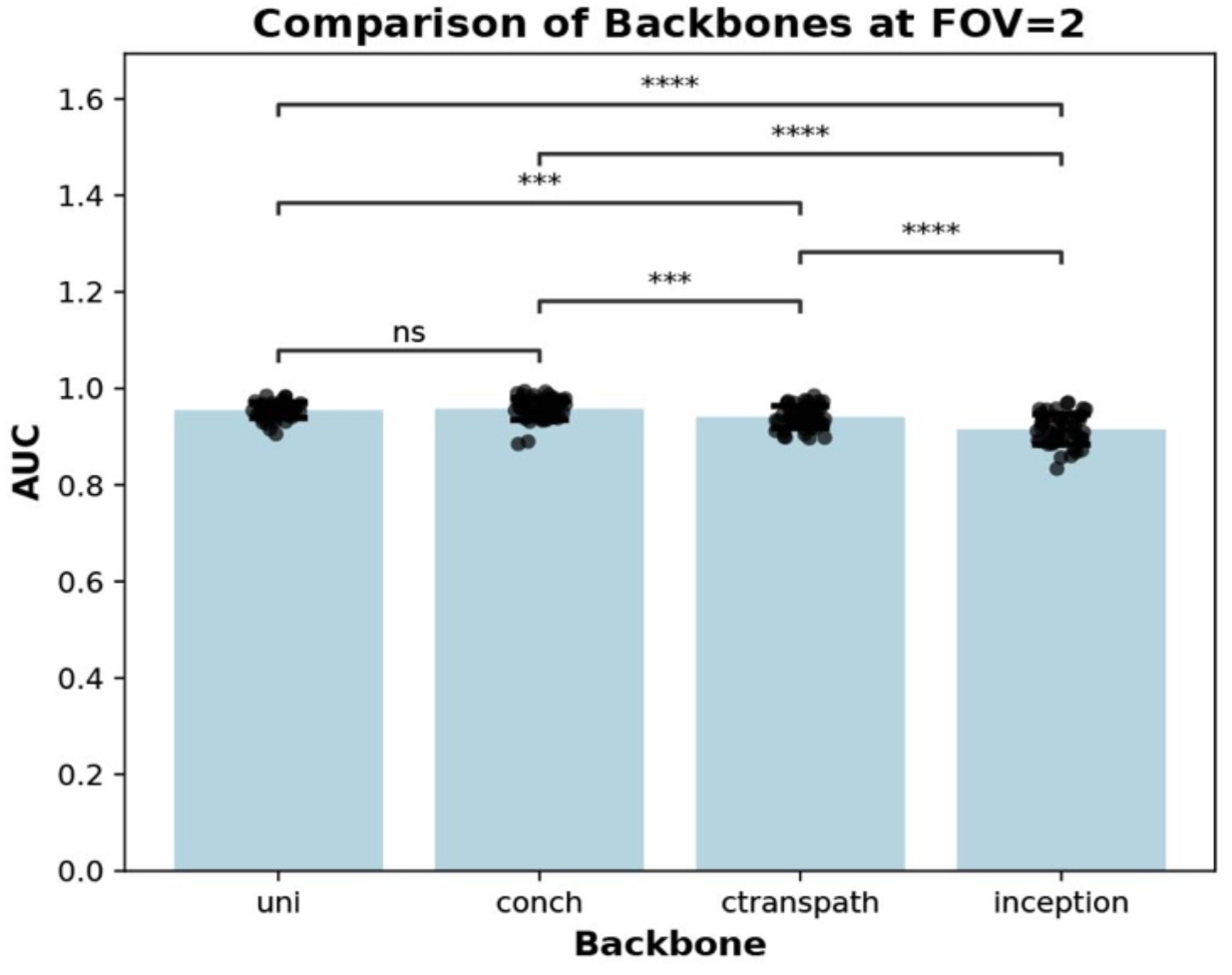
Differences in performance of the various backbones at the FOV 2 for the task of alveolar vs embryonal classification, 100 bootstrapping iterations per 5 folds.

**Supplementary Figure S5:**
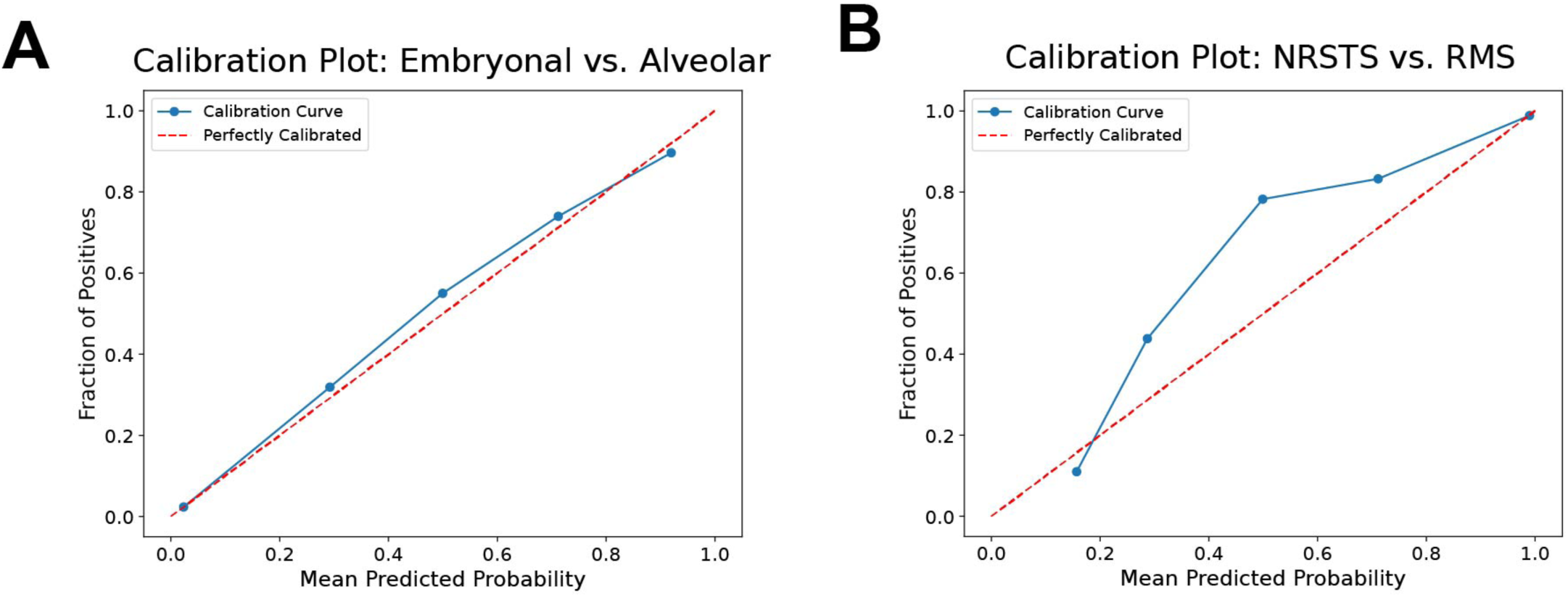
Model calibration plots for the best feature extractor combinations (predictions on all test folds) across 50 iterations for **A,** Embryonal vs. Alveolar and **B,** NRSTS vs. RMS.

**Supplementary Figure S6.**
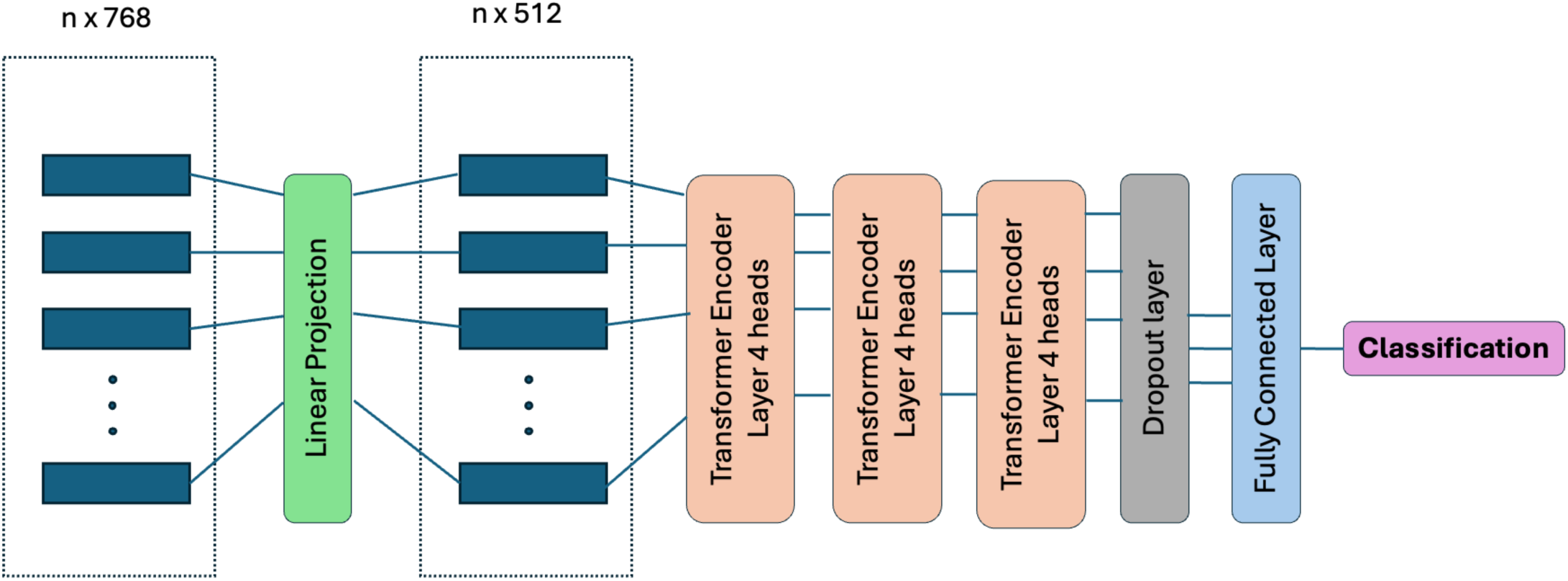
General Transformer architecture used for WSI classification. Initially tiled images must be synthesized and distilled to one output class. All the tiles of an image are linearly projected to reduce the patch dimension to 512. All tiles are passed through three MHSA Transformer encoder layers with 4 heads each. A dropout layer is used to reduce overfitting. Finally, a fully connected layer distills in information from all the patches into a final probability output for each class.

**Supplementary Figure S7.**
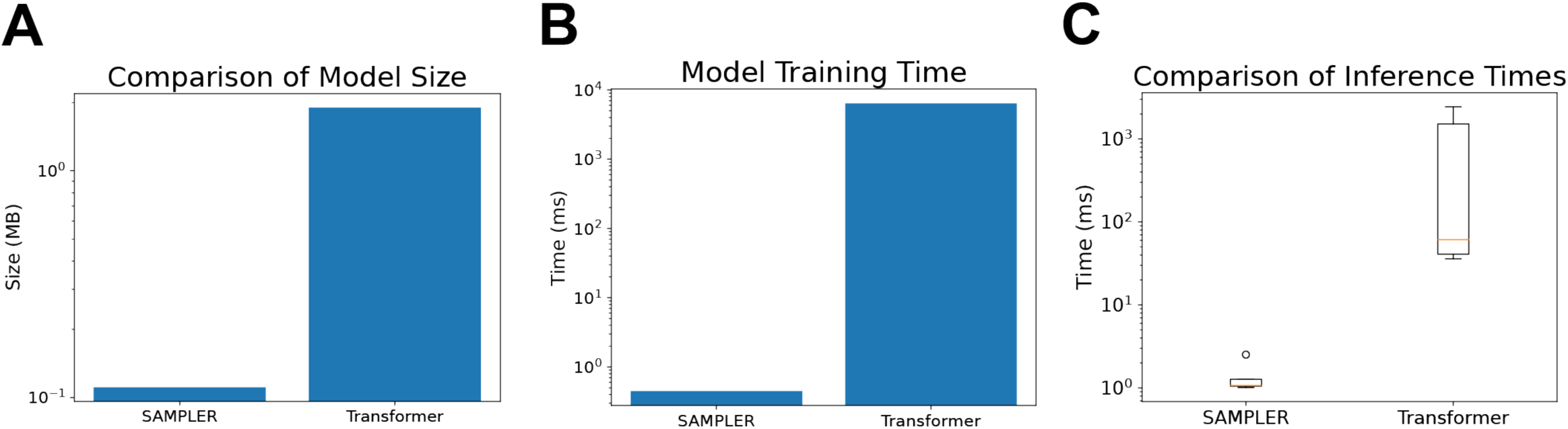
Comparison of runtime and size for SAMPLER vs Transformer models. **A,** Model size difference in megabytes (MB) when saved to disc. **B,** Time to train the model over the full dataset, leaving 5 samples for inference testing. **C,** Comparison of inference times for both models on CPU.

**Supplementary Figure S8.**
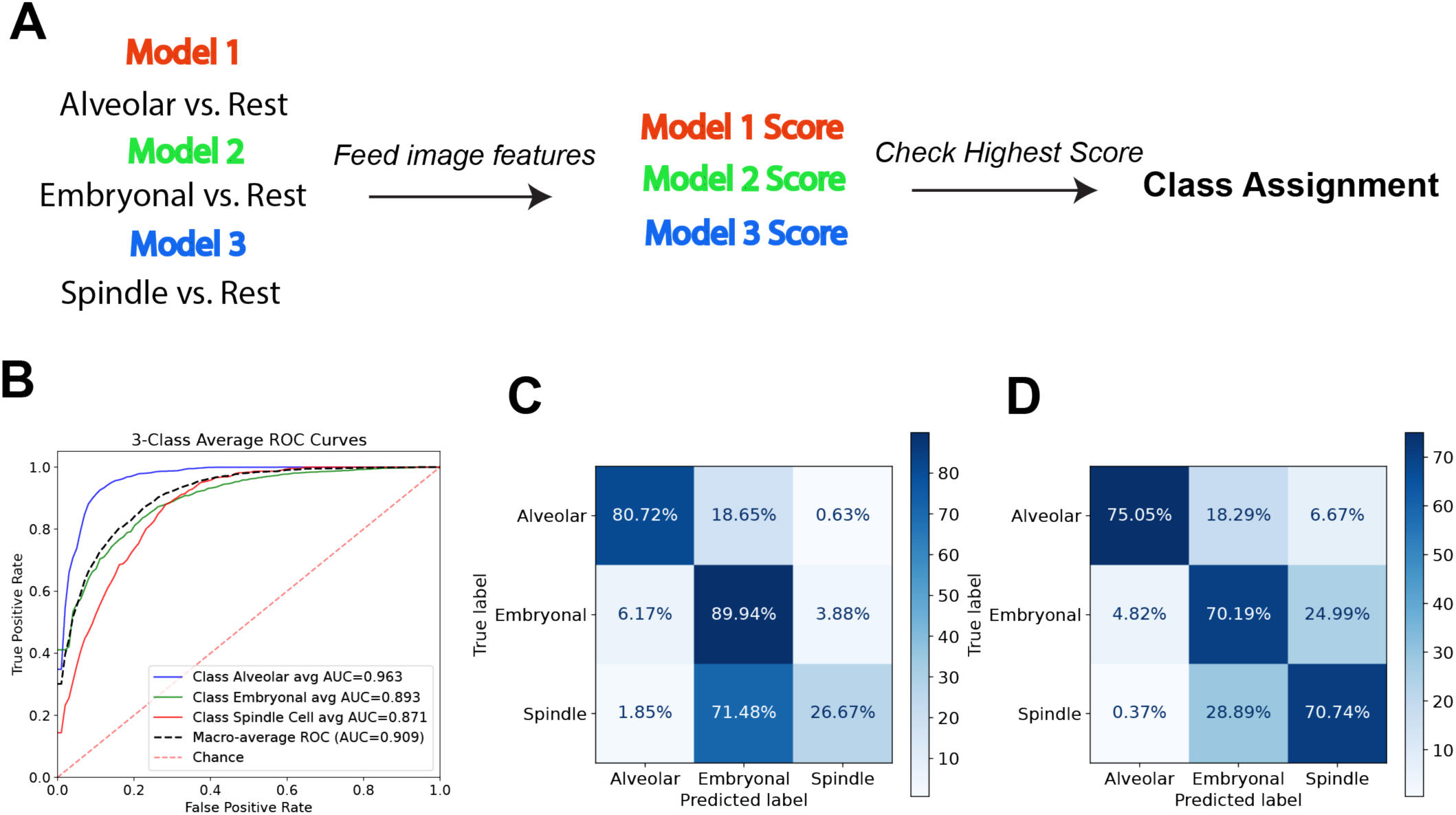
Overview of the OvR based multiclassification approach. Separate one-vs-rest models are trained. **A,** Image features of the test WSI are fed into each model, and the model with the highest prediction score determines the assigned class. **B,** ROC curves for alveolar, embryonal and spindle cell subtypes. **C,** Confusion Matrix showing classification predictions of the 3 subtypes. **D,** Confusion matrix at a lower threshold (0.00023) for more accurate spindle cell classification.

**Supplementary Figure S9.**
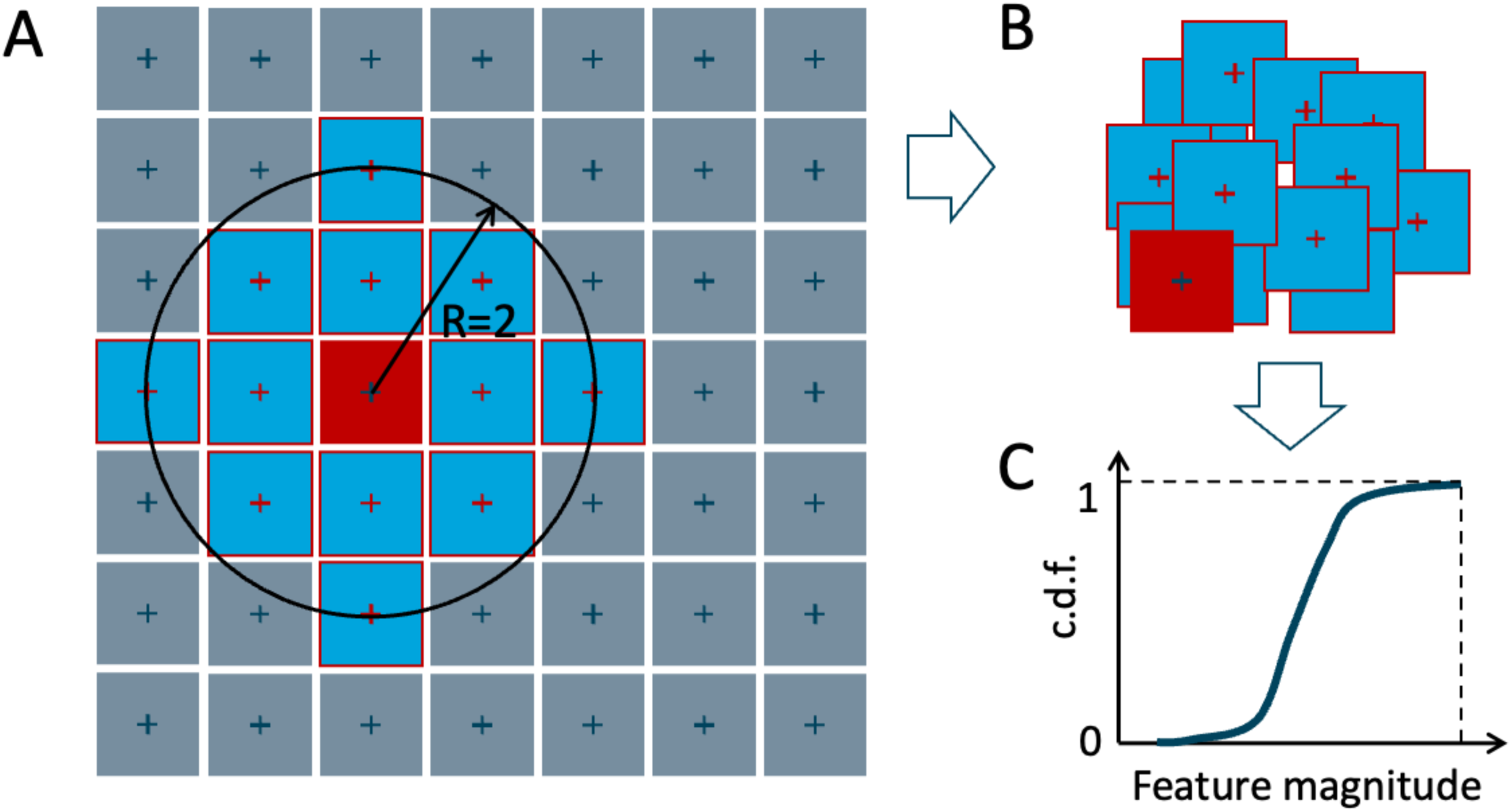
Local SAMPLER spatial inference approach. **A,** Schematic showing how the local neighborhoods for each cell are chosen, here using as an example R=2. **B,** The features for all of the tiles in the neighborhood are then examined together. **C,** For each feature, the c.d.f. is then constructed across all tiles. This creates a representative summary of the features in the neighborhood, which can be compared with the SAMPLER representations of the WSI to find regions of similarity.

## References

1. Florou, V., Nascimento, A. G., Gulia, A. & de Lima Lopes, G. Global Health Perspective in Sarcomas and Other Rare Cancers. Am. Soc. Clin. Oncol. Educ. Book 916–924 (2018) doi:10.1200/EDBK_200589.

2. Children’s Oncology Group’s 2023 Blueprint for Research: Soft Tissue Sarcomas - PMC. https://pmc.ncbi.nlm.nih.gov/articles/PMC10519430/.

3. Grünewald, T. G. et al. Sarcoma treatment in the era of molecular medicine. EMBO Mol. Med. 12, e11131 (2020).

4. Rhabdomyosarcoma: Updates on classification and the necessity of molecular testing beyond immunohistochemistry - ClinicalKey. https://www.clinicalkey.com/#!/content/playContent/1-s2.0-S0046817723002514?returnurl=null&referrer=null.

5. Williamson, D. et al. Fusion Gene–Negative Alveolar Rhabdomyosarcoma Is Clinically and Molecularly Indistinguishable From Embryonal Rhabdomyosarcoma. J. Clin. Oncol. 28, 2151–2158 (2010).

6. Gounder, M. M. et al. Clinical genomic profiling in the management of patients with soft tissue and bone sarcoma. Nat. Commun. 13, 3406 (2022).

7. Lopes, C. D. H. et al. Discordance Between the Initial Diagnosis of Sarcomas and Subsequent Histopathological Revision and Molecular Analyses in a Sarcoma Reference Center in Brazil. JCO Glob. Oncol. e2300431 (2024) doi:10.1200/GO.23.00431.

8. Raut, C. P. et al. High rates of histopathologic discordance in sarcoma with implications for clinical care. J. Clin. Oncol. 29, 10065–10065 (2011).

9. Zhou, J. et al. Integrative deep learning analysis improves colon adenocarcinoma patient stratification at risk for mortality. eBioMedicine 94, 104726 (2023).

10. Zhang, Z. et al. Pathologist-level interpretable whole-slide cancer diagnosis with deep learning. Nat. Mach. Intell. 1, 236–245 (2019).

11. Rubinstein, J. C. et al. Deep learning image analysis quantifies tumor heterogeneity and identifies microsatellite instability in colon cancer. J. Surg. Oncol. 127, 426–433 (2023).

12. Howard, F. M. et al. The impact of site-specific digital histology signatures on deep learning model accuracy and bias. Nat. Commun. 12, 4423 (2021).

13. Milewski, D. et al. Predicting Molecular Subtype and Survival of Rhabdomyosarcoma Patients Using Deep Learning of H&E Images: A Report from the Children’s Oncology Group. Clin. Cancer Res. 29, 364–378 (2023).

14. Frankel, A. O. et al. Machine learning for rhabdomyosarcoma histopathology. Mod. Pathol. 35, 1193–1203 (2022).

15. Zhang, X. et al. Deep Learning of Rhabdomyosarcoma Pathology Images for Classification and Survival Outcome Prediction. Am. J. Pathol. 192, 917–925 (2022).

16. Ho, D. J. et al. Toward Deploying a Deep Learning Model for Diagnosis of Rhabdomyosarcoma. Mod. Pathol. 37, 100421 (2024).

17. Madusanka, N., Jayalath, P., Fernando, D., Yasakethu, L. & Lee, B.-I. Impact of H&E Stain Normalization on Deep Learning Models in Cancer Image Classification: Performance, Complexity, and Trade-Offs. Cancers 15, 4144 (2023).

18. Khan, A. et al. Impact of scanner variability on lymph node segmentation in computational pathology. J. Pathol. Inform. 13, 100127 (2022).

19. Mukashyaka, P., Sheridan, T. B., Foroughi pour, A. & Chuang, J. H. SAMPLER: unsupervised representations for rapid analysis of whole slide tissue images. eBioMedicine 99, 104908 (2024).

20. Vaswani, A., et al. Attention Is All You Need. Preprint at 10.48550/arXiv.1706.03762 (2023).

21. Wagner, S. J. et al. Transformer-based biomarker prediction from colorectal cancer histology: A large-scale multicentric study. Cancer Cell 41, 1650–1661.e4 (2023).

22. Domanskyi, S. et al. Nextflow pipeline for Visium and H&E data from patient-derived xenograft samples. Cell Rep. Methods 4, (2024).

23. Neidlinger, P., et al. Benchmarking foundation models as feature extractors for weakly-supervised computational pathology. Preprint at 10.48550/arXiv.2408.15823 (2024).

24. Szegedy, C., Vanhoucke, V., Ioffe, S., Shlens, J. & Wojna, Z. Rethinking the Inception Architecture for Computer Vision. Preprint at 10.48550/arXiv.1512.00567 (2015).

25. Wang, X. et al. Transformer-based unsupervised contrastive learning for histopathological image classification. Med. Image Anal. 81, 102559 (2022).

26. Chen, R. J. et al. Towards a general-purpose foundation model for computational pathology. Nat. Med. 30, 850–862 (2024).

27. Lu, M. Y. et al. A visual-language foundation model for computational pathology. Nat. Med. 30, 863–874 (2024).

28. Pan, Z., Zhuang, B., Liu, J., He, H. & Cai, J. Scalable Vision Transformers with Hierarchical Pooling. Preprint at 10.48550/arXiv.2103.10619 (2021).

29. Davicioni, E. et al. Molecular Classification of Rhabdomyosarcoma—Genotypic and Phenotypic Determinants of Diagnosis. Am. J. Pathol. 174, 550–564 (2009).

30. Hostein, I. et al. Rhabdomyosarcoma: Value of myogenin expression analysis and molecular testing in diagnosing the alveolar subtype. Cancer 101, 2817–2824 (2004).

31. Grünewald, T. G. P. et al. Ewing sarcoma. Nat. Rev. Dis. Primer 4, 1–22 (2018).

32. Fayzullina, D. et al. Novel Targeted Therapeutic Strategies for Ewing Sarcoma. Cancers 14, 1988 (2022).

33. Yasui, N. et al. Clinicopathologic analysis of spindle cell/sclerosing rhabdomyosarcoma. Pediatr. Blood Cancer 62, 1011–1016 (2015).

34. MemoryFormer: Minimize Transformer Computation by Removing Fully-Connected Layers. https://arxiv.org/html/2411.12992v1.

35. Hrycej, T., Bermeitinger, B. & Handschuh, S. Number of Attention Heads vs Number of Transformer-Encoders in Computer Vision. in Proceedings of the 14th International Joint Conference on Knowledge Discovery, Knowledge Engineering and Knowledge Management 315–321 (2022). doi:10.5220/0011578000003335.

36. Liu, Z., et al. Swin Transformer: Hierarchical Vision Transformer using Shifted Windows. Preprint at 10.48550/arXiv.2103.14030 (2021).

37. Salvi, M. & Molinari, F. Multi-tissue and multi-scale approach for nuclei segmentation in H&E stained images. Biomed. Eng. OnLine 17, 89 (2018).

38. Carrington, A. M. et al. Deep ROC Analysis and AUC as Balanced Average Accuracy, for Improved Classifier Selection, Audit and Explanation. IEEE Trans. Pattern Anal. Mach. Intell. 45, 329–341 (2023).

39. Ahmed, I. et al. Convolutional Neural Network for Histopathological Osteosarcoma Image Classification. Comput. Mater. Contin. 69, 3365–3381 (2021).

40. Gohlke, C. cgohlke/tifffile: v2025.1.10. Zenodo 10.5281/zenodo.14630101 (2025).

41. Senaras, C., Niazi, M. K. K., Lozanski, G. & Gurcan, M. N. DeepFocus: Detection of out-of-focus regions in whole slide digital images using deep learning. PLoS ONE 13, e0205387 (2018).

42. Lu, M. Y. et al. A visual-language foundation model for computational pathology. Nat. Med. 30, 863–874 (2024).

43. Korsunsky, I. et al. Fast, sensitive and accurate integration of single-cell data with Harmony. Nat. Methods 16, 1289–1296 (2019).

